# Longitudinal Metabolomics of Human Plasma Reveals Robust Prognostic Markers of COVID-19 Disease Severity

**DOI:** 10.1101/2021.02.05.21251173

**Authors:** Miriam Sindelar, Ethan Stancliffe, Michaela Schwaiger-Haber, Dhanalakshmi S. Anbukumar, Randy A. Albrecht, Wen-Chun Liu, Kayla Adkins Travis, Adolfo García-Sastre, Leah P. Shriver, Gary J. Patti

## Abstract

There is an urgent need to identify which COVID-19 patients will develop life-threatening illness so that scarce medical resources can be optimally allocated and rapid treatment can be administered early in the disease course, when clinical management is most effective. To aid in the prognostic classification of disease severity, we performed untargeted metabolomics profiling of 341 patients with plasma samples collected at six longitudinal time points. Using the temporal metabolic profiles and machine learning, we then built a predictive model of disease severity. We determined that the levels of 25 metabolites measured at the time of hospital admission successfully predict future disease severity. Through analysis of longitudinal samples, we confirmed that these prognostic markers are directly related to disease progression and that their levels are restored to baseline upon disease recovery. Finally, we validated that these metabolites are also altered in a hamster model of COVID-19. Our results indicate that metabolic changes associated with COVID-19 severity can be effectively used to stratify patients and inform resource allocation during the pandemic.

## Introduction

Coronavirus disease 2019 (COVID-19), which is caused by infection with the novel coronavirus SARS-CoV-2, has led to a global health crisis (Wu et al., 2020b). As of January 2021, more than 100 million cases of COVID-19 have been reported worldwide and resulted in over 2.1 million deaths (2020d). The infection fatality rate of SARS-CoV-2 can be reduced with the appropriate care (e.g., intensive care unit beds, staff, extracorporeal life support, and therapeutics). Such resources are limited, however, and with fewer than five million individuals in the United States fully vaccinated, they continue to be in high demand (2020d). In the United States, one out of five hospitals with an intensive care unit (ICU) has at least 95% of their ICU beds full (Conlen F., 2021) and fewer than 150,000 patient courses of casirivimab and imdevimab monoclonal antibodies have been distributed (2020c). Availability of bamlanivimab, the only other monoclonal antibody that currently has emergency use authorization, has been similarly limited (2020b).

To reduce mortality, patients who develop critical illness from COVID-19 must be treated early in the disease course before the onset of severe symptoms (Kim et al., 2020). Unfortunately, COVID-19 progresses rapidly and it is currently difficult to determine which subset of infected patients will develop life-threatening disease (Kattan et al., 2020). If these patients could be identified, however, then the limited amount of resources available could be optimally allocated to save the greatest number of lives. To this end, the objective of the current study was to identify metabolites in patient plasma that accurately predict life-threatening cases of COVID-19 prior to the onset of severe symptoms.

SARS-CoV-2 is an enveloped, single-stranded positive-sense RNA virus that gains entry into host cells through binding of the viral S protein to the angiotensin-converting enzyme 2 (ACE2) receptor (Hou et al., 2020; Zhang et al., 2020). Multiple studies have established that patients infected with SARS-CoV-2 have metabolic dysregulation, possibly due to immune-triggered inflammation or other changes in host physiology (Fraser et al., 2020; Kimhofer et al., 2020; Overmyer et al., 2020; Shen et al., 2020; Thomas et al., 2020; Wu et al., 2020a). To date, however, unique alterations in metabolites upon SARS-CoV-2 infection have not been validated in large patient cohorts. Moreover, metabolites have not been profiled longitudinally from early after infection through recovery to assess which changes are indicative of disease course.

In this study, we performed untargeted metabolomics profiling on the polar and non-polar fractions of over 700 human plasma samples collected from 341 patients as part of the WU-350 cohort recruited during the first phase of the pandemic in St. Louis, MO. Untargeted metabolomics allows for the unbiased profiling of the human metabolome (Patti et al., 2012) and has been successful at discovering metabolite biomarkers associated with disease pathology (Beger et al., 2016). Using machine learning, we built a predictive model of COVID-19 disease severity based on the metabolic profiles of samples collected from patients at hospital presentation. The model led us to identify 25 unique metabolite biomarkers that were highly predictive of future disease severity. We confirmed that these metabolites were directly related to SARS-CoV-2 infections through comparison to patient demographics, comorbidities, clinical measurements, and longitudinal samples taken from individuals over the course of disease progression. Lastly, we validated that the same biomarkers appeared in an established hamster model of SARS-CoV-2 infection (Chan et al., 2020; Imai et al., 2020; Muñoz-Fontela et al., 2020).

## Results

### Clinical cohort WU-350 – demographics

The clinical cohort presented in this study consisted of 155 female and 186 male participants. Out of the 341 patients, 274 tested positive for SARS-CoV-2 by nasopharyngeal swab PCR and 67 tested negative. The demographic information for the patients is summarized in Table 1.

**Table 1.** Demographics of All Subjects^1,2,3^.

| Parameter | COV- | COV+ | p-value |
| --- | --- | --- | --- |
| n | 67 | 274 |  |
| Gender (M/F) | 28/39 | 158/116 | p=0.0193 |
| Age (yr) | 48 ± 16 | 60 ± 17 | p<0.0001 |
| Race (African American/White/other) | 32/35/0 | 203/66/5 | p<0.0001 |
| Body mass index | 30 ± 8 | 31 ± 9 | p=0.3518 |
| COVID-19-like symptoms |  |  |  |
| Any number of COVID19-like symptoms | 65 | 253 | p=0.1710 |
| Fever | 29 | 129 | p=0.5764 |
| Chills | 13 | 46 | p=0.6120 |
| Conjunctival congestion | 1 | 1 | p=0.2786 |
| Nasal congestion | 7 | 10 | p=0.0219 |
| Headache | 18 | 23 | p<0.0001 |
| Cough | 35 | 148 | p=0.7939 |
| Sore throat | 14 | 23 | p=0.0032 |
| Shortness of breath | 44 | 168 | p=0.5097 |
| Nausea or vomiting | 15 | 41 | p=0.1414 |
| Diarrhea | 11 | 38 | p=0.5939 |
| Myalgia | 16 | 66 | p=0.9717 |
| Fatigue | 21 | 54 | p=0.0393 |
| Loss of taste or smell<sup>4</sup> | 0 | 4 | p=0.3198 |
| No COVID-19-like symptoms<sup>5</sup> | 2 | 21 | p=0.1710 |
| Comorbidities |  |  |  |
| Acute respiratory failure with hypoxia and/or hypercapnia | 15 | 101 | p=0.0250 |
| Chronic kidney disease | 9 | 90 | p=0.0017 |
| Acute renal failure | 1 | 21 | p=0.0653 |
| Diabetes | 16 | 132 | p=0.0003 |
| Cancer (history/current) | 5 (3/2) | 14 (9/5) | p=0.5347 |
| Laboratory Results |  |  |  |
| Hypoxia | 17 | 117 | p=0.0092 |
| Low arterial pH | 5 | 72 | p<0.0001 |
| High arterial pH | 5 | 71 | p=0.0011 |
| Low/High arterial pH | 8 | 92 | p=0.0005 |
| Critically low/high arterial pH | 1 | 35 | p=0.0071 |
| Current smoker | 18 (27%) | 34 (12.4%) | p=0.0032 |
| Hospital admission<sup>6</sup> | 26 (38.8%) | 253 (92.3%) | p<0.0001 |
| ICU admission<sup>6</sup> | 10 (14.9%) | 129 (47.3%) | p<0.0001 |
| Intubation and mechanical ventilation | 4 (6.0%) | 49 (17.9%) | p=0.0154 |
| Deceased | 4 (6.0%) | 52 (19.0%) | p=0.0097 |
| Deceased because of COVID-19 | N/A | 48 (92.3%) | p<0.0001 |

<sup>1</sup> Includes both training and test cohort
<sup>2</sup> Data are presented as mean ± standard deviation, p values of numeric parameters calculated using a 2-tailed Student's t-test with unequal variance, p value of categorical parameters calculated using a chi-square test.
<sup>3</sup> Abbreviations: M – male, F – female, yr – years, B – African American, W – White, O – Other, Y – yes, N – no
<sup>4</sup> CDC guideline symptom was added to the symptom questionnaire late in the study, parameter is not available for the majority of the subjects
<sup>5</sup> SARS-CoV-2 test was routinely administered at presentation at the hospital. The latter was for reasons other than the COVID-19 disease, e.g. accidents, pre-operation tests, regular check-ups, cancer screening, injuries, chest pain
<sup>6</sup> Hospital and/or ICU admission for other reasons than COVID-19 disease symptoms, e.g. accidents, acute respiratory failure due to pneumonia, intentional self-harm, possible heart failure, hypertension, trauma, cancer

Significant differences were observed in several demographic factors for the SARS-CoV-2-positive (COV+) cohort compared to the SARS-CoV-2-negative (COV-) cohort. The age ranges of both the COV+ and COV-cohorts are comparable (Figure S1a). However, the COV+ group has significantly older study participants (p<0.0001). The COV+ group is also enriched for African American, male, and non-smoking individuals. There was no significant difference in the body mass index (BMI) between the two groups (Figure S1b).

Out of 274 COV+ individuals, 253 were admitted to the hospital and 129 of those patients were admitted to the ICU. As expected, the incidence of both factors (hospitalization and ICU admission), were significantly increased in the COV+ cohort. Treatment of severe COVID-19 cases often results in intubation and mechanical ventilation (Goyal et al., 2020). In total, 49 of the COV+ patients required mechanical ventilation, whereas only four COV-individuals required mechanical ventilation. The mortality rate in the COV+ group was 19%, which was significantly higher than in the COV-group (6.0%). A total of 52 COV+ patients died, with 48 of the deaths being attributed to COVID-19 and 4 being attributed to other causes.

Out of 274 COV+ patients, 253 showed at least one COVID-19-related symptom mentioned by the Centers for Disease Control and Prevention (CDC) including fever, chills, conjunctival congestion, nasal congestion, headaches, cough, sore throat, shortness of breath, nausea or vomiting, diarrhea, myalgia, fatigue, and loss of taste or smell (CDC, 2020). The remaining 21 COV+ cases showed none of these symptoms and were classified as COVID-19-asymptomatic. Patients without COVID-19 symptoms received a SARS-CoV-2-test upon presentation at the hospital for reasons unrelated to the pandemic (e.g., accidents, trauma, routine procedures, pre-operation testing, or cancer screening/treatment). Out of the 67 COV-cases, 58 presented with at least one COVID-19-related symptom, while two did not have any symptoms characteristic of COVID-19. The frequency of COVID-19-related symptoms is shown in Table 1, and the distributions across the COV+ and COV-cohorts are depicted in Figure S1c. In both the COV- and COV+ groups, the number of COVID-19 related symptoms reported per individual was comparable. The breakdown of how many symptoms were experienced per individual in both the COV+ and COV-groups is shown in Figure S1d.

Next, we examined the distribution of comorbidities in the presented WU-350 cohort. The incidence of acute respiratory failure with hypoxia and/or hypercapnia, chronic kidney disease (CKD), and diabetes was significantly higher in the COV+ group compared to the COV-group (Table 1). Of the COV+ patients, 43% were hypoxic at some point during their hospitalization, a significantly higher proportion than in the COV-group (25%). Furthermore, 34% of COV+ individuals showed an abnormal arterial pH compared to 12% in the COV-group.

### Study design

Blood was collected from study participants enrolled in the WU-350 study during their initial presentation at the hospital (d0). Further longitudinal samples were collected 3 (d3), 7 (d7), 14 (d14), 28 (d28), and 84 (d84) days after the initial blood collection when possible. However, the collection of longitudinal samples depended on survival of the study participants as well as the participants’ compliance to donate blood samples after being discharged from the hospital. A total of 704 human plasma samples from 341 patients were available for metabolomics profiling, including 324 d0 samples, 165 d3 samples, 111 d7 samples, 54 d14 samples, 31 d28 samples, and 19 d84 samples. All samples were divided into nine randomized sample batches and analyzed by liquid chromatography/mass spectrometry (LC/MS). An extract of the standard reference material SRM 1950 from NIST (National Institute of Standards and Technology, Metabolites in Frozen Human Plasma) was measured repeatedly as a quality control (QC) and blank samples were used to assess background signals. Polar and lipid metabolite fractions were extracted from each sample, and a global metabolomics profile was acquired in both positive and negative ionization modes. Processing of the data led to the putative identification of 235 polar and 472 lipid metabolites based on accurate mass and MS/MS matching. Peak areas were extracted for these 707 metabolites to form the metabolic profile of each patient.

Given that the metabolic profiles were acquired over several months, the combined data showed strong batch effects as demonstrated by the principal component analysis (PCA) in Figure S2a. To remove the variance introduced by the individual batches, but not lose the differentiating biological variance within the research (WU-350) samples, we tested several normalization approaches (Figure S2b) and selected a Combined Batch Correction (ComBat) (Fernández-Albert et al., 2014) approach that outperformed the other common normalization approaches tested (e.g., PQN, unit length, constant sum, quantile, etc.). After normalization, the metabolic profiles retained differences according to sample origin (WU-350, QC, blank) as shown in Figure S2c but no longer clustered based on batch (Figure S2d).

The goal of this study was to find metabolic alterations that are predictive of disease severity in SARS-CoV-2 positive individuals. We used admission to the ICU during disease progression to classify patients as having severe or non-severe disease, as has been done previously (Arunachalam et al., 2020; Petrilli et al., 2020). An ideal biomarker panel would allow an individual presenting at the hospital and receiving a positive SARS-CoV-2-PCR-test result to be screened for metabolic markers associated with severe disease progression to guide the best treatment at the earliest stage of hospitalization. Thus, we grouped the presented COV+ cohort into a non-severe (COV+ non-severe) group that did not require ICU admission and a severe group (COV+ severe) that did require ICU admission. For data interpretation purposes, two study samples were excluded due to a missing SARS-CoV-2-PCR-test result, one sample due to missing clinical information, and 15 samples were excluded as they represented longitudinal samples from COV-individuals. The final patient cohort consisted of 67 COV-cases, 145 COV+ non-severe cases, and 129 COV+ severe cases. Unsupervised analysis of the metabolic profiles for the 324 d0 samples available in our patient cohort demonstrated a clear trend in principal components space that separated COV+ severe, COV+ non-severe, and COV-patients (Figure 1a). Further, several significantly varying metabolites suggested that the metabolic profiles at d0 may indeed be predictive of disease severity. Hierarchical clustering analysis (HCA) of the 54 statistically significant metabolites (p<0.05, Welch’s ANOVA) with an absolute fold change greater than two when compared to the COV-group revealed striking changes in multiple representatives of lipid classes including lysophophatidylcholines (LPCs), phosphatidylcholines (PCs), and triglycerides (TGs). Further, several polar metabolites known to be related to COVID-19 including gluconate (Song et al., 2020) and dimethylguanosine (Migaud et al., 2020) were also significantly altered (Figure 1b).

**Figure 1.**
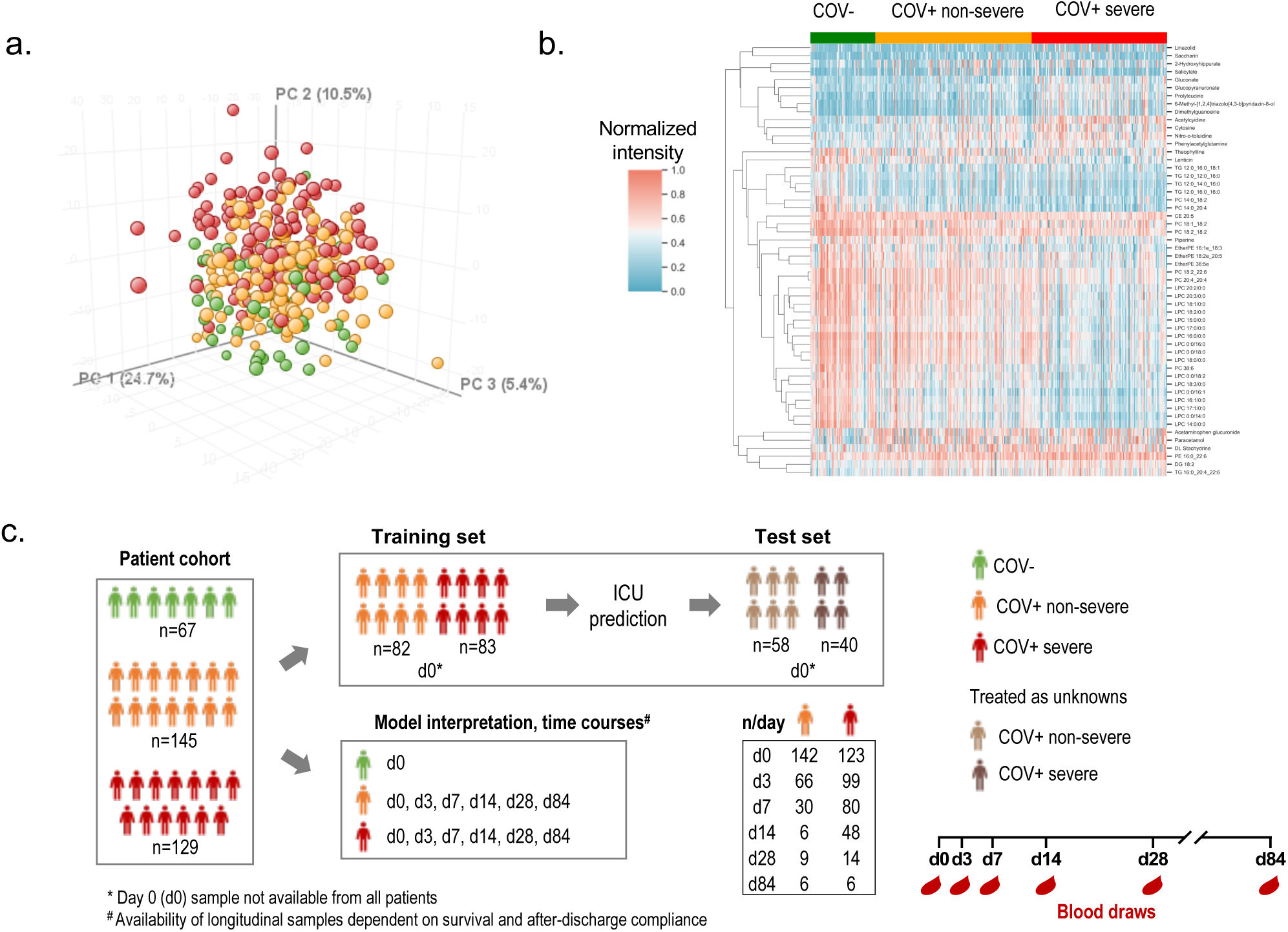
Study design. a) Principal component analysis based on all polar (n=235) and lipid (n=472) metabolites in SARS-CoV-2-negative individuals (COV-, n= 67, green), SARS-CoV-2-positive individuals with non-severe disease (COV+ non-severe, n=142, orange), and SARS-CoV-2-positive individuals with severe disease (COV+ severe, n=123, red) based on the sample provided during presentation at the hospital (d0). b) Hierarchical cluster analysis of metabolic profiles of COV-, COV+ non-severe, and COV+ severe patients at d0. Represented are 54 significantly changing polar and lipid metabolites (p<0.05, Welch’s ANOVA, Benjamini-Hochberg correction). Each column is a sample and each row is a metabolite. c) Human cohort of 341 patients presenting at Barnes Jewish Hospital and Christian Hospital in St. Louis, Missouri. Nasal swab SARS-CoV-2-PCR testing resulted in 67 SARS-CoV-2-negative and 274 SARS-CoV-2-positive participants. The cohort was divided into a training cohort and a test cohort. The study design incorporated 6 blood draws for SARS-CoV-2-positive individuals on days 0 (d0), 3 (d3), 7 (d7), 14 (d14), 28 (d28), and 84 (d84) days after presentation at the hospital.

### Predictive model of COVID-19 disease severity

The global trends in the d0 metabolic profiles visible in the PCA and HCA visualizations prompted us to develop a machine learning (ML) model of disease severity that would predict ICU admission caused by SARS-CoV-2-infection. To make this prediction, we relied on the metabolic signatures in blood plasma at the day of hospital presentation (d0). The 707 metabolites that composed the metabolic profiles served as the predictors for our ML model. To assess predictive power, we split our dataset into two distinct groups: a training set (165 patients) that we used to select, optimize, and train our ML model and a test set (98 patients) that was only used to evaluate the model’s performance (Figure 1c, Table S1, Table S2). Using our training set, we evaluated the efficacy of five ML algorithms with 20-fold cross validation and found that a linear ElasticNet (Zou and Hastie, 2005) regression model was the most effective (Figure S3a). After training the model, we applied it to the patients in the test set and assessed performance by using the area under the receiver operating characteristic curve (AUC). On the test set, we see strong predictive performance (AUC = 0.72) that outperforms a simple model that only uses BMI and age to predict disease severity (Figure 2a) and is significantly more predictive than a random model (Figure 2b, see *Permutation test* in Methods). As further validation, when the trained model was applied to the COV-patients (no COV-patients were in the training set), the mean scores output by the model were lower than those for the COV+ non-severe and the COV+ severe patients in the test set (Figure S3b). This indicates that the model can not only differentiate disease severity but also can distinguish COV+ and COV-patients. We wish to emphasize that PCR is the gold standard to diagnose SARS-CoV-2 infection. As such, we present this result only as confirmation that our model correctly predicts disease severity and not as a diagnostic for viral infection.

**Figure 2.**
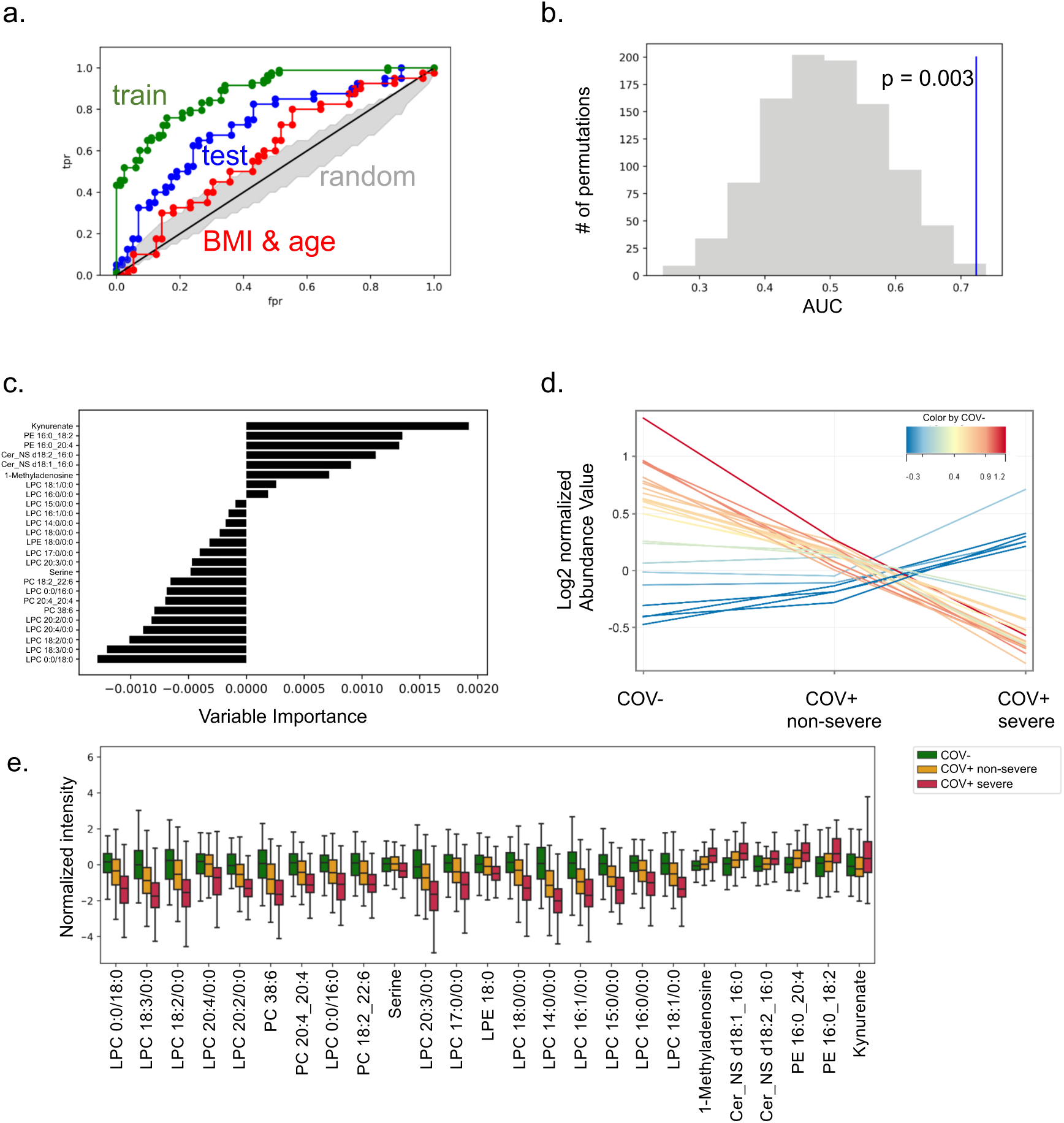
Predicting SARS-COV-2 severity by machine learning. a) Receiver operating characteristic (ROC) curve of prediction model on training set (green) and test set (blue). Random performance is shown in grey. ROC of BMI and age as predictors for severe COVID-19 (red) results in nearly random performance. b) Permutation test results from permuting training set labels and training the model on the permuted data. With every permutation, the area under the ROC curve (AUC) was computed. The histogram shows the distribution of these AUC values for 1000 random permutations. In blue, the model performance on the test set when trained on the non-permuted training data results in an empirical p-value of 0.003. c) Variable importance in reduced ElasticNet prediction model (25 metabolite predictors) for disease severity of SARS-CoV-2-infection in humans. Negative values are predictive of non-severe disease and positive values are predictive of severe disease. Variable importance is after the model is trained on the complete dataset. d) Profile plot of the normalized signal abundance of 25 prediction model metabolites grouped into COV- (control, n=67), COV+ non-severe (n=142), and COV+ severe (n=123). e) Boxplots showing predictor metabolite intensities in the COV-, COV+ severe, and COV+ non-severe groups. Box limits represent the quartiles of each sample group. Whiskers are drawn to 1.5x of the inter-quartile range.

We next sought to interpret which metabolites were most salient to the model’s predictions. First, we computed the variable importance of the model when trained on the complete dataset, which found 93 metabolites that contributed to the model’s predictions. Among this group of 93 compounds were metabolites that have been previously implicated in SARS-CoV-2 infection such as bilirubin, kynurenate, nicotinamide, creatinine, LPCs, and others (Shen et al., 2020; Song et al., 2020; Thomas et al., 2020; Wu et al., 2020a). The mean intensity of each metabolite in the COV-, COV+ non-severe, and COV+ severe groups can be seen in Figure S4. Next, we aimed to assess the robustness of the metabolites selected by the ML model. We used bootstrap resampling of our training dataset to construct confidence intervals for the variable importance of each of the 707 metabolites profiled (Mendez et al., 2020). The analysis led to the identification of 25 metabolites that significantly contributed to the model’s fit. The structural identities of these metabolites were rigorously confirmed (see Methods). Strikingly, 14 of the 25 metabolites are LPCs. Using this reduced predictor set, we re-trained and re-optimized our ElasticNet model on the training set and assessed the predictive power of these 25 metabolites on our test set.

Using only these 25 metabolites resulted in nearly an identical AUC to when the full set of metabolites was used (AUC = 0.70) and still performed better than a random model or a model that used only BMI and age as predictors (Figure S5). The variable importance of these 25 metabolites when trained on the entire dataset is shown in Figure 2c. The mean intensity of the 25 metabolites in the COV-, COV+ non-severe, and COV+ severe groups is shown in Figure 2d and Figure S6. All LPCs and PCs that contributed to the model, as well as serine, presented a downward trend of signal abundance with disease severity. Conversely, the other polar metabolites, (kynurenate and 1-methyladenosine) and two phosphatidylethanolamines (PEs), exhibited an upward trend in signal intensity (Figure 2e).

### Demographics, laboratory values, comorbidities, and COVID-19 severity

After evaluating the efficacy of the ML model, we wished to deduce the relationship of our 25 robust metabolite predictors to COVID-19 disease severity. We examined whether these metabolites were reflective of an underlying condition, risk factor for severe disease, or related to the disease progression of COVID-19. We addressed the former by asking whether any of the 25 metabolites correlated with demographic factors, laboratory values, or individual patient comorbidities available for the patient cohort. A comparison of the COV+ non-severe and severe groups identified several significantly different parameters (Table 2). The COV+ severe group is significantly biased towards patients with advanced age, however, the age ranges in both groups are comparable (Figure 3a). There was no significant difference in BMI (Figure 3b), but we note that there was variability in BMI for both patient groups. CO_2_ levels were not significantly altered between groups (Figure 3c), with values mostly being in the normal range. In contrast, there were significantly increased levels of the inflammatory marker C-reactive protein (Figure 3d). D-dimer, absolute neutrophil count, and neutrophil % were also increased (Figure 3e-g).

**Table 2.** Demographics, Comorbidities and Lab values of SARS-CoV-2-infected individuals with d0 sample available^1,2,3^.

| Parameter | COV+ non-severe | COV+ severe | p-value |
| --- | --- | --- | --- |
| n | 142 | 123 |  |
| Gender (M/F) | 75/67 | 77/46 | p=0.1082 |
| Age (yr) | 55 ± 17 | 66 ± 15 | p<0.0001 |
| Age range (yr) | 19.2 - 92.7 | 19.3 - 90.8 |  |
| Race (African American/White/other) | 109/30/3 | 88/35/0 | p=0.1987 |
| BMI | 32 ± 9 | 30 ± 10 | p=0.1107 |
| Current smoker | 24 | 11 | p=0.0564 |
| Deceased | 3 | 47 | p<0.0001 |
| Deceased due to COVID-19 | 2 | 44 | p<0.0001 |
| **Comorbidities** |  |  |  |
| Acute respiratory failure | 27 | 68 | p<0.0001 |
| Chronic kidney disease | 40 | 49 | p=0.0449 |
| Acute renal failure | 9 | 12 | p=0.3043 |
| Diabetes | 59 | 68 | p=0.0256 |
| Cancer (history/current) | 5 (3/2) | 8 (5/4) | p=0.2622 |
| **Laboratory Results** |  |  |  |
| Hypoxia | 34 | 76 | p<0.0001 |
| Low arterial pH | 4 | 62 | p<0.0001 |
| High arterial pH | 4 | 61 | p<0.0001 |
| Low/High arterial pH | 6 | 80 | p<0.0001 |
| Extreme pH | 0 | 33 | p<0.0001 |
| C-reactive protein (mg/L) | 67.8 ± 65.45 (n=89) | 154.6 ± 110.9 (n=101) | p<0.0001 |
| D-dimer (ng/mL FEU) | 2614 ± 6839 (n=91) | 5895 ± 10682 (n=103) | p=0.108 |
| Neutrophil absolute (K/cumm) | 4.806 ± 2.635 (n=135) | 7.644 ± 5.602 (n=117) | p<0.0001 |
| Neutrophil (%) | 66.95 ± 12.03 (n=135) | 78.01 ± 12.10 (n=117) | p<0.0001 |
| CO<sub>2</sub>, Total (mmol/L) | 24.86 ± 3.78 (n=138) | 24.10 ± 4.17 (n=122) | p=0.1287 |

<sup>1</sup> Includes both training and test cohort
<sup>2</sup> Data are presented as mean ± standard deviation, p values of numeric parameters calculated using a 2-tailed Student's t-test with unequal variance, p value of categorical parameters calculated using a chi-square test.
<sup>3</sup> Abbreviations: M – male, F – female, yr – years, B – African American, W – White, O – Other, Y – yes, N – no

**Figure 3.**
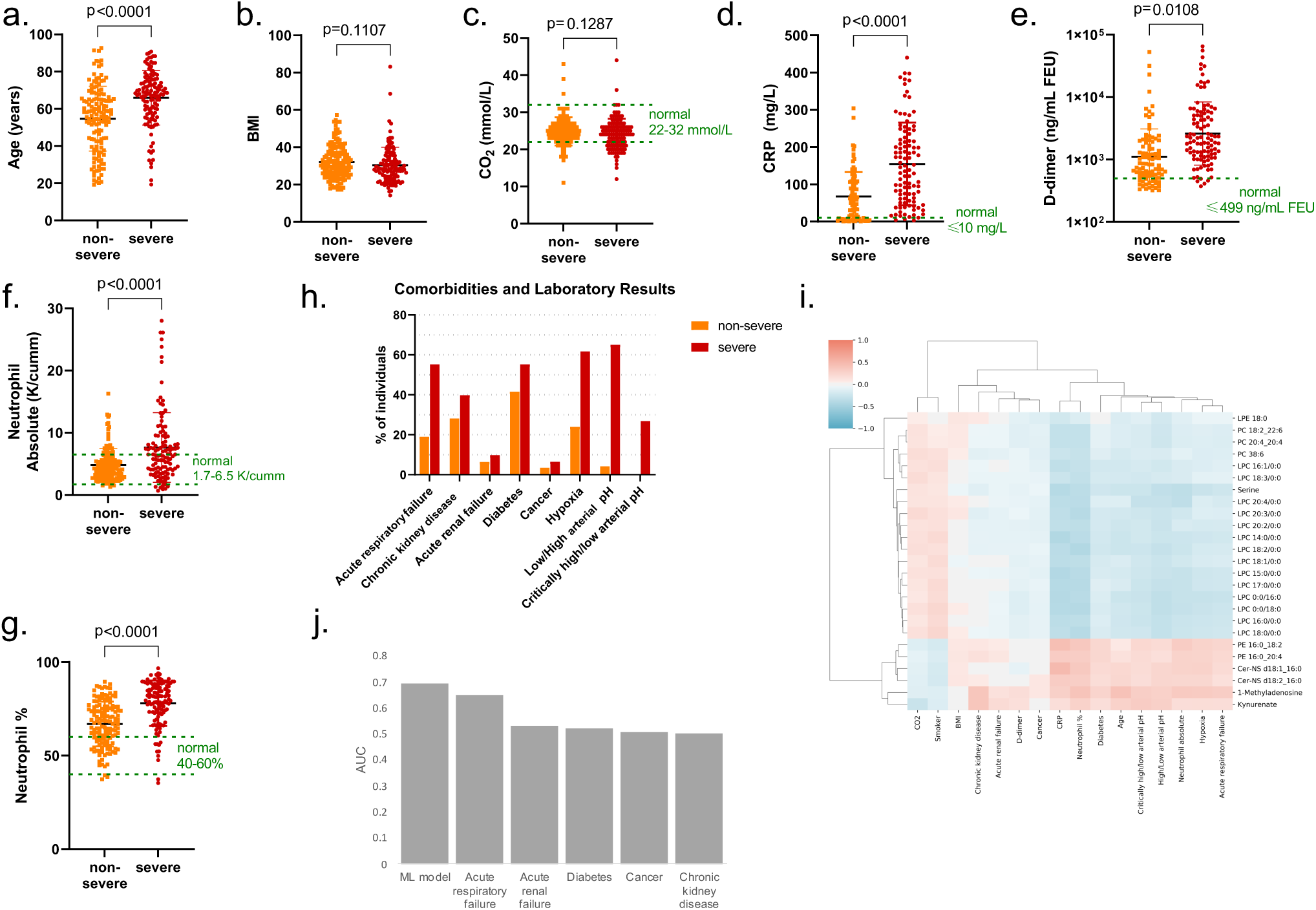
COV+ patient parameters. Demographics, comorbidities, and laboratory values of SARS-CoV-2-positive cases grouped by disease severity (non-severe, severe) for age (a), BMI (b), CO_2_ (c), C-reactive protein (d), D-dimer (e), absolute neutrophil levels (f), and neutrophil percentage (g). Statistical significance was assessed with a 2-tailed Student’s t-test with unequal variance for data shown in (a-g). h) proportion of COV+ severe and non-severe patients with particular comorbidities and laboratory test results. i) Pearson correlation of listed demographic/laboratory results/comorbidities with abundances of the predictor metabolites. j) Area under the ROC curve (AUC) values for patient comorbidities and the ML model when predicting disease severity on the test set patients.

These data indicate more severe inflammation in the COV+ severe group compared to the non-severe group and are consistent with reports from previous studies (Ahmed et al., 2020; Luo et al., 2020; Thomas et al., 2020). Neutrophil recruitment has also been shown to be dysregulated in severe COVID-19 disease (Liao et al., 2020; Park and Lee, 2020; Yang et al., 2020; Zhou et al., 2020).

Next, because specific comorbidities increase the risk of having a severe case of COVID-19 (Jain and Yuan, 2020; Petrilli et al., 2020; Smith et al., 2020), we also asked which co-morbidities are enriched in the COV+ severe group compared to the COV+ non-severe group (Figure 3h, Table 2). The COV+ severe patients had a significantly greater proportion of individuals suffering from acute respiratory failure, CKD, and/or diabetes. The number of individuals with cancer (or a history of cancer) and acute renal failure was not significantly different between the groups. Further, laboratory tests showed an increased proportion of individuals having hypoxia and abnormal arterial pH in the COV+ severe group compared to COV+ non-severe patients. Critically high/low pH values were only observed in the COV+ severe group. We note that timestamps for laboratory tests and measurements were not available for the patient cohort due to HIPPA privacy regulations. As such, these tests and measurements could have been performed at any point during an individual’s hospital stay.

Considering the number of significant associations in the patient parameters between COV+ severe and non-severe patients, we wanted to check whether any of our predictor metabolites significantly correlated with the clinical data. To that end, we computed the Pearson correlation (Benesty et al., 2009) (for continuous parameters) or the point biserial correlation (Tate, 1954) (for binary parameters) between each predictor metabolite and patient parameter (Figure 3i). The analysis did not reveal any strong correlations between patient parameters and our predictor metabolites. The only significant but moderate correlation with age was with 1-methyladenosine (r=0.4), which was also correlated weakly with CKD (r=0.37) and neutrophil percentage (r=0.39). Significant but weak correlations (r=0.37) were observed for kynurenate and CKD, which has been described previously (Gagnebin et al., 2020). Further, C-reactive protein and neutrophil percentages have a moderate positive correlation with the ceramide (Cer) Cer-NS d18:1_16:0 (r=0.47) and PE 16:0_18:2 (r=0.43). Both the C-reactive protein values and the neutrophil percentages are weakly to moderately negatively correlated with most of the LPCs and serine levels (r = [−0.4, −0.31]), indicating that the reduction of LPCs and serine is concomitant with the immune response to SARS-CoV-2 infection (Mudd et al., 2020). Notably, the majority of our predictor metabolites had only weak or insignificant correlations with the comorbidities or patient parameters.

We next sought to assess the predictive power of our ML model (when trained on the training set) relative to the predictive power of the patient comorbidities. Thus, for each patient comorbidity we computed the AUC when predicting disease severity based on the comorbidity status for each patient in the test set (Figure 3j). For all evaluated comorbidities, the model achieves a higher AUC. Taken together, these results suggest that our predictor metabolites are indeed relevant to the pathogenesis of SARS-CoV-2 infection and not merely markers of other risk factors.

### Longitudinal progression of predictor metabolites

To give further confidence that our predictor metabolites are associated with COVID-19 pathogenesis, we next aimed to determine how the levels of these metabolites changed over the course of disease progression. First, we considered the portion of the COV+ severe cohort that survived SARS-CoV-2-infection. We sought to determine the temporal behavior of their metabolic profiles as patients reach peak disease severity and after recovery. Accordingly, we compared the longitudinal metabolite abundances from individuals who had severe disease but survived and were discharged from the hospital. We compared their initial d0 plasma sample with the sample taken closest to the day of ICU admission and the last sample provided by the patient at or after hospital discharge. For several LPCs and one PC, a V-shaped trend was observed (Figure 4a). After the initial sample (d0), the level of these metabolites dropped further as the disease worsened but then began to restore during recovery. The reverse trend was observed for Cer-NS d18:1_16:0. Its levels significantly increased until the patients were admitted to the ICU. However, the levels sharply dropped to below the initial d0 levels in the final sample obtained.

**Figure 4.**
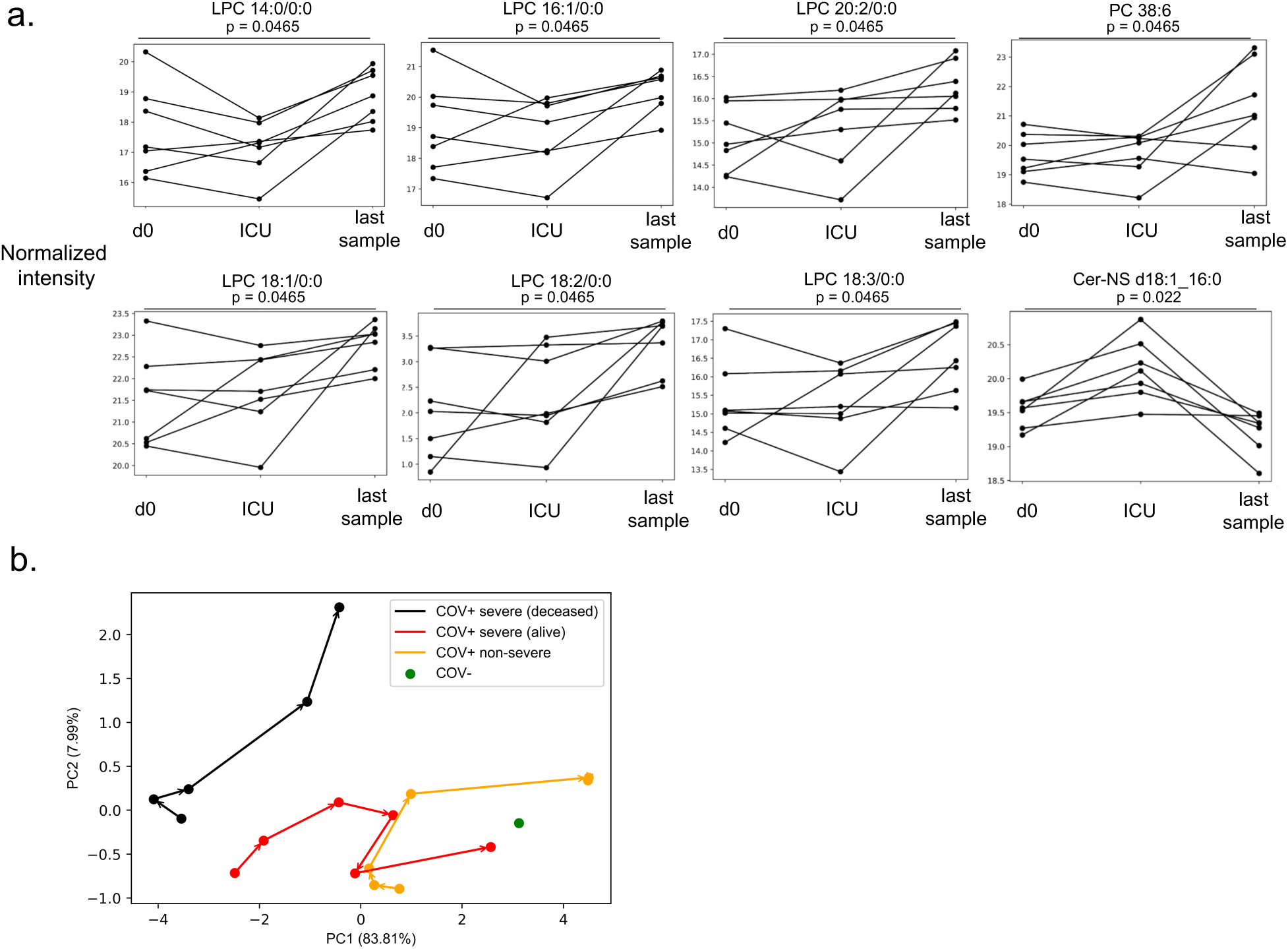
Course of disease progression. a) Prediction model metabolites that significantly vary in intensity as a function of disease progression for SARS-CoV-2 positive patients surviving severe disease (COV+ severe). d0 denotes the first sample after hospital admission, ICU denotes the sample collected closest to ICU admission, and the last sample is the final sample collected for the patient. Only patients where these time points were distinct samples were used. Statistical significance was assessed by using a repeated measures one-way ANOVA with Benjamini-Hochberg correction. b) Principal components analysis showing the trajectory of the mean metabolic profile of the 25 predictor metabolites in COV+ non-severe patients (orange), surviving COV+ severe patients (red), and deceased COV+ severe patients (black). No d84 samples were available for deceased COV+ severe patients. The last two points for COV+ non-severe patients overlap. In green, the mean d0 metabolic profile of COV-patients is shown. The surviving COV+ patient profiles approach the d0 COV-profile by d84.

These pronounced longitudinal trends in surviving COV+ severe patients raised the question of how the trajectory of disease progression (as marked by our predictor metabolites) differed among COV+ non-severe patients, surviving COV+ severe patients, and deceased COV+ severe patients. Further, we wished to compare the end points in these groups to the COV-d0 patients. We constructed representative metabolite profiles for the groups by using the 25 predictor metabolites at each of the study time points (d0, d3, d7, d14, d28, and d84) and performed a principal component analysis that enabled the trajectory of each group to be drawn out in two dimensions (Figure 4b). Strikingly, the analysis revealed three distinct trajectories with starting points that trended with disease severity. The groups then followed a common trajectory through d14, after which deceased and surviving COV+ severe patients diverge, and COV+ non-severe patients rapidly progress to the end of the trajectory that is constant for d28 and d84. For both surviving COV+ severe patients and COV+ non-severe patients, the endpoint is close to that of the d0 COV-patients. However, COV+ non-severe patients reach this point faster. Conversely, for deceased COV+ patients, after d14, the metabolic profile moves further away from the COV-profile staying relatively constant in principal component one (explaining 84% of the variance) but increasing away from COV-in principal component two (explaining 8% of variance). No d84 samples were available for deceased COV+ severe patients. We next examined the individual metabolite levels within the four groups at each time point. The deceased COV+ severe patients show the same direction of dysregulation across the predictor metabolites as the surviving COV+ severe patients, but the magnitude of the perturbation is increased. Unlike the other groups, these deceased patients show no recovery throughout the disease progression (Figure S7).

Next, we sought to compare the longitudinal progression of the predictor metabolites between the surviving COV+ patients. In the COV+ severe group, the LPC levels increased over the course of 84 days to levels that are comparable to the COV-group (FC=1, Figure 5a). In the COV+ non-severe group, the LPC levels recovered faster, and, at day 28, an overcompensation occurred resulting in higher LPC levels than in the COV-group (FC=1, Figure 5b). In total, 22 out of the 25 predictor metabolites showed a significant change (p<0.05, Welch’s ANOVA) across the longitudinal timepoints in the COV+ severe group (Figure 5c). All 14 LPCs significantly increased over time, as well as lysophosphatidylethanolamine (LPE) 18:0, PC 38:6, PC 20:4_20:4, and PC 18:2_22:6. Kynurenate, Cer-NS d18:1_16:0, Cer-NS d18:2_16:0, and PE 16:0_20:4 showed a decreasing trend after initially being increased compared to the d0 sample of the COV-group. Due to lower sample numbers, the COV+ non-severe group had only 11 metabolites that showed a significant trend (p<0.05, Welch’s ANOVA, Figure 5d). These 11 metabolites are composed of 8 LPCs, Cer-NS d18:1_16:0, PC 38:6, and LPE 18:0.

**Figure 5.**
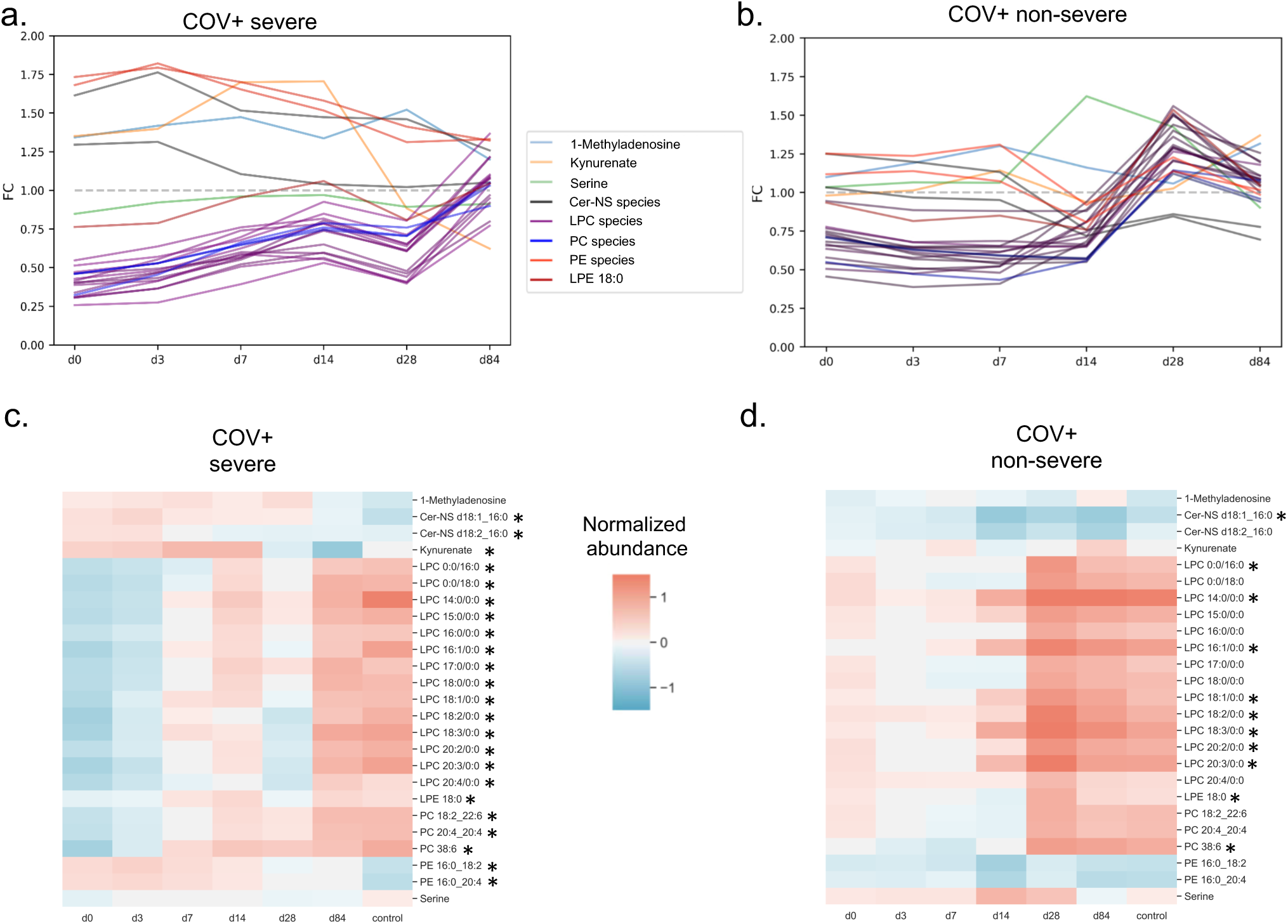
Longitudinal trends in COV+ patients. Changes in plasma levels of the 25 predictor metabolites over the course of the SARS-CoV-2-infection (d0 through d84). a) Profile plot of the mean predictor metabolite intensities relative to d0 COV-samples (n=67, grey) in SARS-CoV-2-positive individuals with severe COVID-19-disease (n=123, COV+ severe) who survived and were discharged from the hospital. b) Profile plot of the mean predictor metabolite intensities in SARS-CoV-2-positive individuals with non-severe disease (n=142, COV+ non-severe). c-d) Heatmaps showing relative mean intensity of predictor metabolites in longitudinal profiles of COV+ severe patients (c) or COV+ non-severe patients (d). The mean COV-d0 profiles are included as the control for reference. * indicates a p-value < 0.05. Statistical significance was assessed using a one-way Welch’s ANOVA with Benjamini-Hochberg correction.

### Syrian Hamster Model Confirms Metabolite Changes in COVID-19 Disease

Lastly, we aimed to validate that the trends observed for the predictor metabolites in the patient samples also appeared in an established animal model of SARS-CoV-2 infection (Chan et al., 2020; Imai et al., 2020). Syrian hamsters have been found to be susceptible to SARS-CoV-2 infection, with the virus mainly replicating in the upper and lower respiratory tract of intranasally challenged animals for approximately six days post-infection. The animals also show signs of disease characterized by body weight lost and pathological lung inflammation. We obtained plasma samples from golden Syrian hamsters that were intranasally inoculated with SARS-CoV-2, influenza virus, or nasally treated with saline solution as a mock infection. Relative to the body weights of the mock hamsters, hamsters infected with SARS-CoV-2 experienced significant bodyweight loss (approximately 15%) while hamsters infected with influenza virus did not lose body weight, which is consistent with a previous report (Iwatsuki-Horimoto et al., 2018). After 2, 4, 6, and 14 days (d2, d4, d6, and d14) post-infection, plasma was harvested from the SARS-CoV-2 and influenza virus-infected hamsters (Figure 6a). For the mock group, plasma was harvested on days 4 and 14 relative to the infection timeline. All plasma samples were subjected to the same LC/MS workflow as described above. Of our 25 metabolite predictors, all but PC 20:4_20:4 was detected in the hamster plasma. We compared samples from the three groups (SARS-CoV-2, influenza, and mock) harvested on d4, when the disease was fully established (Chan et al., 2020; Imai et al., 2020). Figure 6b summarizes the significantly changing predictor metabolites (p<0.05, Welch’s ANOVA) across the three groups. The LPCs showed the same trend as what was observed in the human samples (i.e., a significant depression when compared to the control group). For all significantly varying metabolites, with the exception of PE 16:2_20:4, infection with the influenza virus showed a similar trend of dysregulation away from control samples but a very different magnitude compared to a SARS-CoV-2 infection. This is consistent with lower rates of body weight loss in influenza virus-infected hamsters as compared to those infected with SARS-CoV-2. Finally, we wanted to determine whether the predictor metabolites in hamsters showed a similar recovery trend over 14 days of infection to what we observed in human patients over a period of 84 days of infection. Indeed, there is a similar trend in the SARS-CoV-2 infected hamsters (Figure 6c) as in the human COV+ samples when compared to the d4 control (mock) samples. LPC levels dropped significantly on d4 and slowly recovered towards the control levels on d14. By comparison, in the influenza virus-infected group, levels of most metabolites approached that of the control group more rapidly (Figure 6d).

**Figure 6.**
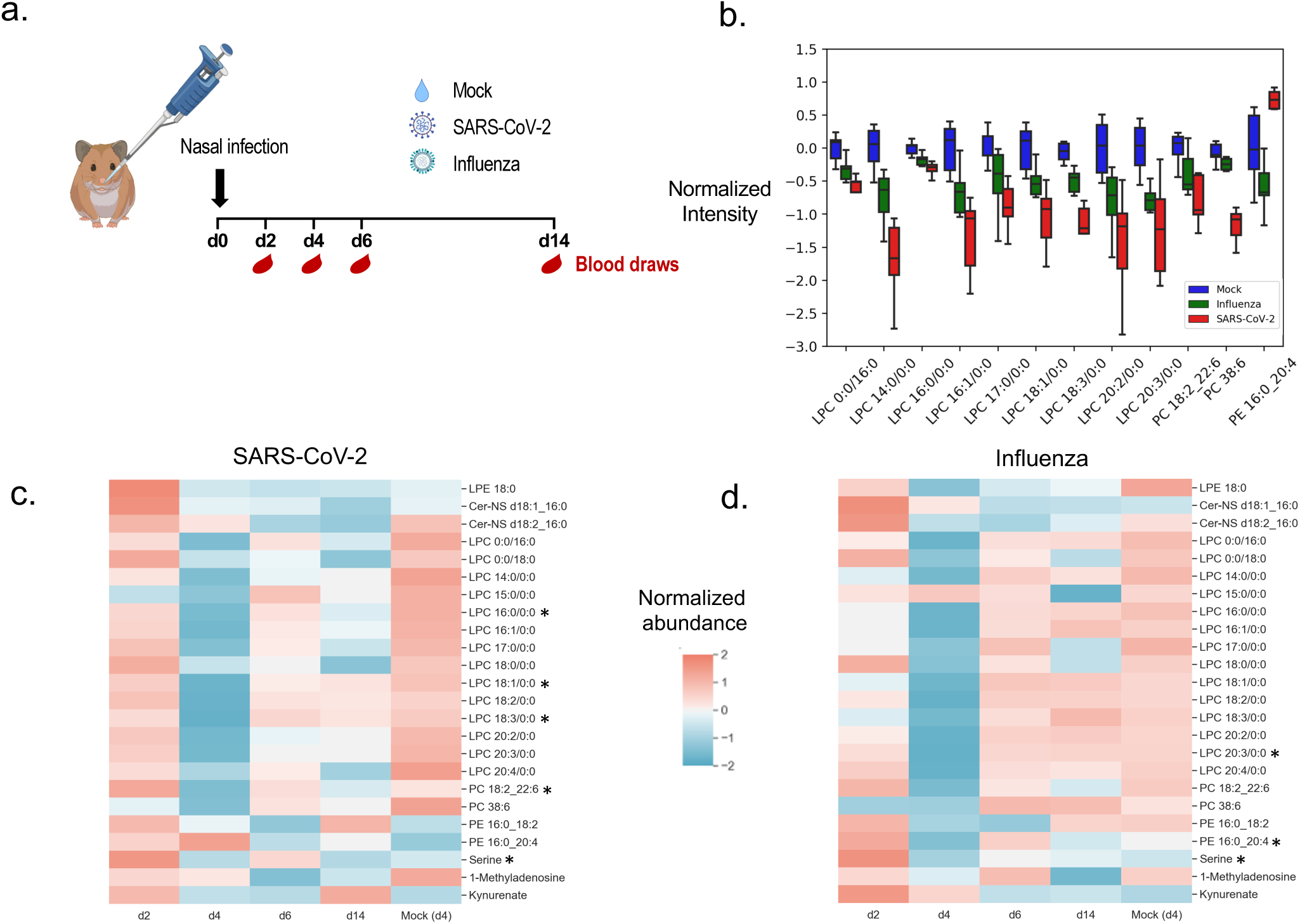
Syrian Hamster model confirms SARS-CoV-2-dependent metabolite changes. a) Experimental design of Syrian hamster model. Hamsters (n=3-6 per group) were infected through intranasal installation of SARS-CoV-2 (1e5 PFU), influenza virus (1e5 PFU), or nasally treated with a saline solution (mock) on day 0 (d0). Blood was drawn 2, 4, 6, and 14 days (d2, d4, d6, d14) post-infection. Nasal washes were performed on day 1, 3, 5, 7, and 9 post-infection. b) Comparing metabolite intensity between hamsters infected with influenza (n=6), SARS-CoV-2 (n=5), and mock (n=6) on d4 shows many of the predictor metabolites are significantly altered in the hamster model (p<0.05, Welch’s ANOVA). Box limits represent the quartiles of each sample group. Whiskers are drawn to 1.5x of the inter-quartile range. c-d) Metabolite changes during disease progression in SARS-CoV-2 (c) and influenza (d) infected animals show a faster recovery for influenza infected animals. All groups are n=6 with the exception of SARS-CoV-2 hamsters at d2 (n=3) and d4 (n=5). * indicates a p-value less than 0.05. Statistical significance was assessed with a 2-tailed Student’s t-test with unequal variance between d2 and d4 samples. All values were corrected with the Benjamini-Hochberg procedure.

## Discussion

The current study sought to predict COVID-19 disease severity based on the metabolic profiles of human plasma samples obtained early in the disease course, prior to the onset of critical illness. We applied untargeted metabolomics to profile a patient cohort of 341 individuals, which amounted to over 700 study samples in addition to QC and method blanks. In these samples, we putatively identified 235 polar metabolites and 472 lipid metabolite species. Using these metabolites, we applied machine learning techniques to build a predictive model that can accurately classify a patient’s disease severity from their day 0 metabolic profile obtained at the time of initial hospital admission. This differentiating power may be critical in the coming months as SARS-CoV-2 infections continue to rise and hospital resources for treating severe disease become increasingly more limited. Due to insufficient availability, for example, casirivimab and imdevimab are currently only indicated for the treatment of patients who are at high risk of progressing to severe COVID-19 (2020a). At this time, risk assessment is based on BMI and age (FDA, 2020). Even though we see a significant difference between the age of the non-severe and severe disease group (p<0.0001), the results of this study show that risk assessment based on our 25 predictor metabolites is more reliable than age and BMI and therefore provides a better metric for resource allocation.

Our linear ElasticNet model is relatively simple compared to other popular ML models, including artificial neural networks, support vector machines (SVM), or ensemble based approaches such as random forest (RF) that have been applied to metabolomics datasets previously (Mendez et al., 2019). However, linear models can be easily interpreted and provide robust performance (Mendez et al., 2019; Zou and Hastie, 2005). Indeed, a previous study (Overmyer et al., 2020) successfully used an ElasticNet model to predict disease severity from a multi-omic dataset. A limitation of the ElasticNet approach is that it is a linear model and most biological systems are innately non-linear. Other studies have used non-linear ML models such as RF to predict disease severity from metabolomics, lipidomics, and/or proteomics profiles (Fraser et al., 2020; Shen et al., 2020). Although these studies found higher AUC scores than that of our model, they used considerably smaller patient cohorts than what our model was trained and evaluated on. When we tested non-linear models (RF and SVM), we found worse cross-validated performance than ElasticNet (see Figure S3a). Another challenge we faced in building a model of disease severity is that the size of our study required normalizing metabolic profiles acquired in multiple batches. We demonstrated that ComBat normalization was able to remove the variance resulting from these batch effects. In removing this variance, however, true biological variation was undoubtedly removed. Despite these limitations, our model still accurately predicted patient disease severity.

Interpretation of our model led us to identify 25 robust predictor metabolites whose identities were rigorously confirmed. Using this reduced predictor set, we were able to retrain our model and found similarly strong predictive ability. Our large sample size that included longitudinal measurements of patient plasma and collection of patient metadata (laboratory values, comorbidities, and demographics) allowed us to uniquely validate the relationship of these 25 metabolites to COVID-19 disease severity. Further, we confirmed the relevance of these metabolites to the pathology associated with SARS-CoV-2 infection by using an established animal model of COVID-19. It is important to point out that the 25 metabolites we discovered to predict disease severity can be readily measured by using targeted methods on triple quadrupole mass spectrometers that are widely available in most clinical laboratories. Thus, the test we present here to assess the risk of a severe case of COVID-19 does not require intensive computation or untargeted metabolomics, making it immediately applicable to most hospitals in the United States.

## Methods

### Study design

Over the period of March to August of 2020, blood specimens of 341 individuals who presented at Barnes Jewish Hospital or Christian Hospital located in Saint Louis, Missouri, USA were collected. Inclusion criteria were a physician-ordered SARS-CoV-2-PCR test with a positive or negative outcome, availability of gender and age information, and an age greater than 18. Informed consent was obtained from all study participants. Samples were collected at the time of enrollment (d0), which was during or immediately following presentation at the hospital, and 3, 7, 14, 28, or 84 days post hospital presentation. Clinically relevant medical information (e.g., patient-reported symptoms, date of symptom-onset, age, race, and BMI) was collected at the time of enrollment from the subject, their legally authorized representative, or the medical record.

### Metabolomics sample preparation

Participant plasma, which had been stored at −80 °C upon collection, was thawed on ice. A 50 µL aliquot was transferred onto the solid-phase-extraction (SPE)-system CAPTIVA-EMR Lipid 96-wellplate (Agilent Technologies) before addition of 250 µL of acetonitrile containing 1% formic acid (v/v) and 10 µM internal standard (consisting of uniformly ^13^C and ^15^N labeled amino acids from Cambridge Isotope Laboratories, Inc). The samples were mixed for 1 min at 360 rpm on an orbital shaker at room temperature prior to a 10 min incubation period at 4 °C. Afterwards, 200 µL 80% acetonitrile in water (v/v) were added to the samples. The samples were mixed on an orbital shaker (360 rpm) for an additional 10 min at room temperature. The samples were then eluted into a 96-deepwell collection plate by centrifugation (10 min, 57 x *g*, 4 °C followed by 2 min, 1000 x *g*, 4 °C). Polar eluates were stored at −80 °C until the day of LC/MS analysis.

The SPE-plates were then washed twice with 500 µL 80% acetonitrile in water (v/v). Lipids still bound to the SPE-material were then released into a second elution plate, in two elution steps applying 2x 500 µL 1:1 methyl *tert*-butyl ether:methanol (v/v) onto the SPE cartridge and centrifuging for 2 min at 1000 g and 4 °C. The combined eluates were dried under a stream of nitrogen (Biotage SPE Dry Evaporation System) at room temperature and reconstituted with 100 µL 1:1 2-propanol:methanol (v/v) prior to LC/MS analysis.

Hamster plasma samples were diluted 1:4 with methanol (v/v), vortexed for 30 seconds and incubated at −20°C for 2 hours. Samples were centrifuged for 10 minutes at 13,500 x *g* at 4°C and supernatant was transferred to a new centrifuge tube, concentrated, and stored at −80°C until reconstitution as described above.

### LC/MS analysis of polar metabolites

An aliquot of 2 µL of polar metabolite extract was subjected to LC/MS analysis by using an Agilent 1290 Infinity II liquid-chromatography (LC) system coupled to an Agilent 6540 Quadrupole-Time-of-Flight (Q-TOF) mass spectrometer with a dual Agilent Jet Stream electrospray ionization source. Polar metabolites were separated on a SeQuant® ZIC®-pHILIC column (100 x 2.1 mm, 5 µm, polymer, Merck-Millipore) including a ZIC®-pHILIC guard column (2.1 mm x 20 mm, 5 µm). The column compartment temperature was maintained at 40 ° C and the flow rate was set to 250 µL×min^-1^. The mobile phases consisted of A: 95% water, 5% acetonitrile, 20 mM ammonium bicarbonate, 0.1% ammonium hydroxide solution (25% ammonia in water), 2.5 µM medronic acid, and B: 95% acetonitrile, 5% water, 2.5 µM medronic acid. The following linear gradient was applied: 0 to 1 min, 90% B; 12 min, 35% B; 12.5 to 14.5 min, 25% B; 15 min, 90% B followed by a re-equilibration phase of 4 min at 400 µL×min^-1^ and 2 min at 250 µL×min^-1^. Metabolites were detected in positive and negative ion mode with the following source parameters: gas temperature 200 °C, drying gas flow 10 L×min^-1^, nebulizer pressure 44 psi, sheath gas temperature 300°C, sheath gas flow 12 L×min^-1^, VCap 3000 V, nozzle voltage 2000 V, Fragmentor 100 V, Skimmer 65 V, Oct 1 RF Vpp 750 V, and m/z range 50-1700. Data were acquired under continuous reference mass correction at *m/z* 121.0509 and 922.0890 for positive ion mode and *m/z* 119.0363 and 966.0007 for negative ion mode. Samples were randomized prior to analysis. In addition, a quality control sample was injected after every 12^th^ sample to monitor signal stability of the instrument.

### LC/MS analysis of lipid metabolites

An aliquot of 2 µL of lipid extract was subjected to LC/MS analysis by using an Agilent 1290 Infinity II LC-system coupled to an Agilent 6545 Q-TOF mass spectrometer with a dual Agilent Jet Stream electrospray ionization source. Lipids were separated on an Acquity UPLC® HSS T3 column (2.1 x 150 mm, 1.8 µm) including an Acquity UPLC® HSS T3 VanGuard Pre-Column (2.1 x 5mm, 1.8 µm) at a temperature of 60 °C and a flow rate of 250 µL×min^-1^. The mobile phases consisted of A: 60% acetonitrile, 40% water, 0.1% formic acid, 10 mM ammonium formate, 2.5 µM medronic acid, and B: 90% 2-propanol, 10% acetonitrile, 0.1% formic acid, 10 mM ammonium formate (dissolved in 1 mL water). The following linear gradient was used: 0-2 min, 30% B; 17 min, 75% B; 20 min, 85%; 23-26 min, 100% B; 26, 30% B followed by a re-equilibration phase of 5 min.

Lipids were detected in positive and negative ion mode with the following source parameters: gas temperature 250 °C, drying gas flow 11 L×min^-1^, nebulizer pressure 35 psi, sheath gas temperature 300 °C, sheath gas flow 12 L×min^-1^, VCap 3000 V, nozzle voltage 500 V, Fragmentor 160 V, Skimmer 65 V, Oct 1 RF Vpp 750 V, and m/z range 50-1700. Data were acquired under continuous reference mass correction at *m/z* 121.0509 and 922.0890 in positive ion mode and *m/z* 119.0363 and 966.0007 in negative ion mode. Samples were randomized before analysis. In addition, a quality control sample was injected after every 12^th^ sample to monitor signal stability of the instrument.

### Data preprocessing and normalization

Polar metabolite identifications were supported by matching the retention time, accurate mass, and MS/MS fragmentation data to our in-house retention time and MS/MS library created from authentic reference standards (Mass Spectrometry Metabolite Library supplied by IROA Technologies, Millipore Sigma, St. Louis, MO, USA) and online MS/MS libraries (Human Metabolome Database (HMDB, https://hmdb.ca, (Wishart et al., 2018)), Mass Bank of North America (MoNA, https://mona.fiehnlab.ucdavis.edu/, (Horai et al., 2010)), and mzCloud (https://mzcloud.org). Lipid iterative MS/MS data were annotated with the Agilent Lipid Annotator software. All data files were then analyzed in Skyline (Version 20.1.0.155) to obtain peak areas. *m/z* values of the metabolite and lipid target lists obtained from the metabolite identification workflow, which had at least an MS/MS match to an online library, were extracted under consideration of retention times.

Due to the risk of handling plasma samples from SARS-CoV-2 positive patients and not knowing how many batches of samples we would receive, we refrained from preparing a pooled sample and instead used the NIST SRM 1950 plasma reference material as quality control (QC) sample in each batch. The QC sample was injected after every 12^th^ sample. After peak area extraction, batch effects were observed in the research samples (see Figure S2a). The research samples and QC data were used to test typical batch normalization methods (see Figure S2b) including constant sum, unit length, scale, percentile shift, minimum-maximum, PQN, quantile and ComBat correction used in metabolomics (Chong et al., 2018; Di Guida et al., 2016; Fernández-Albert et al., 2014; Ghosh, 2017; Johnson et al., 2007). In Figure S2b, the variance remaining in the research samples normalized to the variance in the QC samples is shown for each method. The higher this ratio, the more variance remains in the research samples and the more batch derived variance in the QC samples is reduced. ComBat correction outperformed the other batch correction approaches tested using this metric. After correction, samples are well clustered according to sample type (WU-350, QC, blank) as shown in Figure S2c. Importantly, within the research samples, there is no clustering by batch (see Figure S2d).

### Animal Studies

All studies were performed at Mount Sinai School of Medicine. Outbred female LVG golden Syrian hamsters were sourced from Charles River Laboratories (Kingston, NY). The hamsters were anesthetized by intraperitoneal injection of a mixture of ketamine and xylazine prior to intranasal inoculation with 0.1 mL of 1e5 plaque-forming units (PFU) of SARS-CoV-2 (WA-1) or H1N1 influenza A virus (A/California/04/2009). On day 2, 4, 6, and 14 days post-infection, 3-6 anesthetized hamsters per infection group were euthanized by exsanguination followed by intracardiac injection of veterinary euthanasia solution (SleepAway; Fort Dodge). Plasma samples were treated by exposure to germicidal UV-C light.

### Study Approval

Portions of the human study relevant to Barnes Jewish Hospital, Christian Hospital, and Washington University were reviewed and approved by the Washington University in Saint Louis Institutional Review Board (WU-350 study approval #202003085, and plasma metabolomics study approval #202004204). All animal studies were approved by the Institutional Care and Use Committee at Mount Sinai School of Medicine, following the humane care and use guidelines set by the institution.

### Machine Learning

Samples were split into two distinct cohorts for training and testing the ML model. D0 COV+ patient samples within batches 1-6 made up the training set and d0 COV+ patient samples from batches 7 through 9 made up the test set. Training and tests sets were treated independently except for batch normalization which was carried out for all patients (including samples collected after d0 and COV-samples) together. Demographics of both training and tests sets are available in Table S1 and Table S2.

Model selection was based on 20-fold cross validation of the training set. Five different ML models: logistic regression, ElasticNet linear regression, partial least squares discriminant analysis (PLSDA), support vector machine (SVM), and random forest were selected for consideration based on interpretability and previous studies (Fraser et al., 2020; Lalmuanawma et al., 2020; Mendez et al., 2019; Shen et al., 2020). Hyperparameters of all models and feature selection strategies were optimized using 20-fold cross validation and a grid search. Two separate feature selection strategies were tested: a correlation-based approach and a statistic-based approach. In the correlation-based approach, the Pearson correlation was computed between each metabolite’s intensity and the disease severity. Then, the top X % of metabolites sorted by absolute correlation were taken as the predictors for the ML model. In the statistic-based approach, a student’s t-test was performed to assess the statistical significance of the differences in each metabolite’s intensity between COV+ severe and COV+ non-severe patients. Absolute fold-change and p-value cutoffs were used to select metabolites. Performance was assessed with the area under the receiver operating characteristic curve (AUC). After optimization, ElasticNet regression achieved the highest AUC on the cross validated training dataset. The ElasticNet model is given below in Equation 1 where X is matrix of metabolic profiles (# of samples x # of metabolites), b is the bias term, y is the sample labels (0 = COV+ non-severe, 1 = COV+ severe), w is the weight of each metabolite to the model prediction, α is the weight of the regularization, and ρ is the mixing parameter between the l1 and l2 norm regularization.

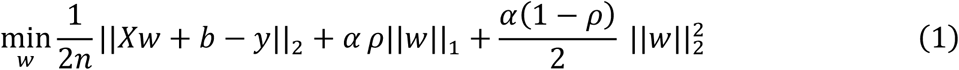

After optimization, the correlation-based feature selection was used taking the top 33% most correlated metabolites with model hyperparameters α = 10.0 and ρ = 0.0. In the reduced predictor model, no feature selection was performed and model hyperparameters α = 1.0 and ρ = 0.0 were used.

The variable importance of each metabolite in the ElasticNet model is easily computed from the optimized weights, w. To normalize for the different abundances of the metabolites, each weight was normalized by the median abundance of the metabolite across all samples. The more positive the variable importance, the more predictive that metabolite is to severe disease. The more negative the variable importance, the more predictive the metabolite is to non-severe disease. To find the metabolites that significantly contribute to the model fit, the training dataset was resampled with replacement 10,000 times. At each iteration, the ElasticNet model was trained and the variable importance was calculated. After the iterations were complete, the 95% confidence interval of the variable importance was calculated for each metabolite using the 2.5 and 97.5 percentiles. If this interval included zero, the metabolite did not significantly contribute to the model fit.

All ML analyses were carried out using Python (v3.7) with extensive use of the packages SciPy (v1.4.1) (Virtanen et al., 2020a) and Scikit-learn (v0.23.1) (Pedregosa et al., 2011).

### Code availability

Custom code used to perform the ML analyses is available on GitHub (https://github.com/e-stan/covid_19_analysis)

### Data availability

The raw LC/MS data as well as the processed metabolic profiles and their corresponding deidentified metadata for the human and animal samples will be made publicly available on the Metabolomics Workbench repository.

### Statistical analysis

All statistical analyses were performed using the SciPy (v1.4.1) (Virtanen et al., 2020b) and statsmodels (v0.11.1) (Seabold and Perktold, 2010) Python packages and with the Mass Profiler Professional Software (Agilent Technologies, v15.5). All p-values were corrected for multiple hypothesis testing using the Benjamini-Hochberg procedure (Benjamini and Hochberg, 1995).

### Permutation test

To assess the significance of the model fit and compare the predictive power to what is expected from random chance, we performed a permutation test. After the feature selection and model hyperparameters were optimized, the training dataset labels were permuted, and the model was retrained on the permuted data. Then, the performance of this model was assessed on the non-permuted test set and the AUC was computed. This process was repeated 1,000 times. The empirical p-value was computed by calculating the percentage of the 1,000 permutations that achieved an AUC higher than that of the model’s performance when trained on non-permuted data.

### Confirming metabolite identities of predictor metabolites

The identities of the 25 predictor metabolites were rigorously confirmed with authentic standards. For the polar compounds, authentic standards were purchased to not only match MS/MS but also retention times for identification. For lipids, one or two standards per lipid class were matched to an authentic standard to compare MS/MS spectra and retention times. PCs were identified based on *m/z* and the two characteristic fragments 184.0733 and 86.0964 in positive ionization mode. For PCs where no peaks for the acyl-chains were observed, only the sum composition can be given. LPE 18:0 was matched to its authentic standard based on retention time and MS/MS spectra. PEs were identified based on the neutral loss of phosphorylethanolamine (141.0191) in positive mode. The fatty acyl composition could be derived from the spectra, but no differentiation of regioisomers was possible, as was the case for ceramides. To denote regiospecificity, metabolites whose regioisomers could be differentiated have their acyl-chains are separated with a “/” while those that could not have a “_”. Cer-NS d18:1_16:0 was matched to its authentic standard. Cer-NS d18:2_16:0 matched the MS/MS library spectrum and eluted slightly before Cer-NS d18:1_16:0 as expected due to having one less double bond. LPCs were identified based on MS/MS spectral matches. Standards were available for LPC 14:0/0:0 and LPC 18:1/0:0. Their retention times were used as a reference for the other LPCs. The two regioisomers of LPCs (sn1 and sn2) were separated by liquid chromatography, with the sn1 isomer eluting later. They are also distinguished by their MS/MS spectra. 1-acyl-LPC (sn1) shows two main fragments (*m/z* 184.0733 and 104.1070), whereas the 2-acyl-LPC (sn2) has a more pronounced 184.0733 fragment. The 104.1070 fragment (choline) has been previously reported as being more abundant from LPCs with the fatty acid chain in the sn1 position from the sodium adducts when studying the lysophospholipid regioisomers (Han and Gross, 1996). We note that sn2 LPCs can be converted to sn1 during sample preparation, and our sample preparation was not dedicated to preserve those isomers (Koistinen et al., 2015; Okudaira et al., 2014).

### Acquiring MS/MS data

MS/MS spectra for polar metabolites were acquired on an Orbitrap ID-X Tribrid mass spectrometer (Thermo Scientific). A Vanquish Horizon UHPLC system, with the same chromatographic conditions as described in the Methods, was interfaced with the mass spectrometer via electrospray ionization in both positive and negative mode with a spray voltage of 3.5 and 2.8 kV, respectively. The RF lens value was 35%. Data were acquired in data dependent acquisition (DDA) mode using the built-in deep scan option (AcquireX) with a mass range of 67-900 *m/z*. MS/MS scans were acquired at 15K resolution on a NIST SRM 1950 plasma sample from and 4 individual samples from d0, d3, d7, and d14 in both positive and negative polarity with different collision energies in the range of 20 NCE to 50 NCE for HCD and 30 NCE for CID to maximize identifications.

MS/MS spectra for polar metabolites and lipids were acquired using an iterative approach in the MassHunter Acquisition Software (Version 10.1.48, Agilent Technologies) on an Agilent 6540 and 6545 QTOF respectively. The same source settings as for MS1 data acquisition were used. MS/MS spectra were acquired at a scan rate of 3 spectra/s with different intensity thresholds and collision energies of 10, 20, and 40 V to increase identification rates.

## Acknowledgements

This work was supported by funding from the National Institutes of Health grants R24OD024624 (G.J.P.) and R35ES2028365 (G.J.P.). This study utilized samples obtained from the Washington University School of Medicine’s COVID-19 biorepository, which is supported by: the Barnes-Jewish Hospital Foundation; the Siteman Cancer Center grant P30 CA091842 from the National Cancer Institute of the National Institutes of Health; and the Washington University Institute of Clinical and Translational Sciences grant UL1TR002345 from the National Center for Advancing Translational Sciences of the National Institutes of Health. The content is solely the responsibility of the authors and does not necessarily represent the view of the NIH. For their work in the development and maintenance of the COVID-19 biorepository, we would like to thank Jane O’Halloran, MD, PhD; Charles Goss, PhD, and Phillip Mudd, MD, PhD. This work was also partly supported by CRIP (Center for Research for Influenza Pathogenesis), a NIAID supported Center of Excellence for Influenza Research and Surveillance (CEIRS, contract HHSN272201400008C; WCL, RAA, and AGS); by NIAID grant U19AI135972; by NCI grant U54CA260560; by supplements to NIAID grants U19AI135972, U19AI142733 and DoD grant W81XWH-20-1-0270; by the Defense Advanced Research Projects Agency (HR0011-19-2-0020); by the generous support of the JPB Foundation and the Open Philanthropy Project (research grant 2020-215611 (5384); and by anonymous donors (A.G.S.).

## Author contributions

M.S. planned the metabolomics study, executed the sample preparation, acquired and analyzed the LC/MS and MS/MS data, did statistical analysis, prepared figures, and wrote the manuscript. E.S. analyzed the data, did the statistical analysis, developed the prediction model, prepared figures, and wrote the manuscript. M.S.-H. executed the sample preparation, acquired and analyzed the LC/MS and MS/MS data, prepared figures, and contributed to writing the manuscript. D.S.A. planned the metabolomics study and collected and managed demographic and clinical information. R.A., W.-C.L., K.A.T., L.P.S and A.G.S. planned the hamster study and collected the samples and prepared the hamster samples for metabolomics. G.J.P. planned the study, helped design experiments, and supervised all aspects of the research. All authors contributed to revising the manuscript.

## Competing interests

The AG-S laboratory has received research support from Pfizer, Pharmamar, Blade Therapeutics, Avimex, Dynavax, Kenall Manufacturing, ImmunityBio, Nanocomposix, Senhwa Biosciences and 7Hills Pharma. AG-S has consulting agreements for the following companies involving cash and/or stock: Vivaldi Biosciences, Contrafect, 7 Hills Pharma, Avimex, Vaxalto, Accurius and Esperovax. G.J.P. is a scientific advisor for Cambridge Isotope Laboratories. All other authors declare no conflicts of interest.

***All human and animal studies have been approved by the appropriate ethics committee and have therefore been performed in accordance with the ethical standards laid down in the 1964 Declaration of Helsinki and its later amendments***.

## Notes

### Author Declarations

The human study relevant to Barnes Jewish Hospital, Christian Hospital, and Washington University were reviewed and approved by the Washington University in Saint Louis Institutional Review Board (WU-350 study approval #202003085, and plasma metabolomics study approval #202004204). All animal studies were approved by the Institutional Care and Use Committee at Mount Sinai School of Medicine, following the humane care and use guidelines set by the institution.

