## supporting information for "Longitudinal Metabolomics of Human Plasma Reveals Robust Prognostic Markers of COVID-19 Disease Severity"

[1]Department of Chemistry, Washington University, St. Louis, MO
[2]Department of Medicine, Washington University, St. Louis, MO
[3]These authors contributed equally
[4]These authors contributed equally
[5]Department of Microbiology, Icahn School of Medicine at Mount Sinai, New York City, NY
[6]Global Health and Emerging Pathogens Institute, Icahn School of Medicine at Mount Sinai, New York City, NY
[7]Current affiliation: Biomedical Translation Research Center, Academia Sinica, Taipei, 11571, Taiwan
[8]Department of Chemistry, University of Akron, Akron, OH
[9]Department of Medicine, Division of Infectious Diseases, Icahn School of Medicine at Mount Sinai, New York City, NY
[10]The Tisch Cancer Institute, Icahn School of Medicine at Mount Sinai, New York City, NY
[11]Siteman Cancer Center, Washington University, St. Louis, MO

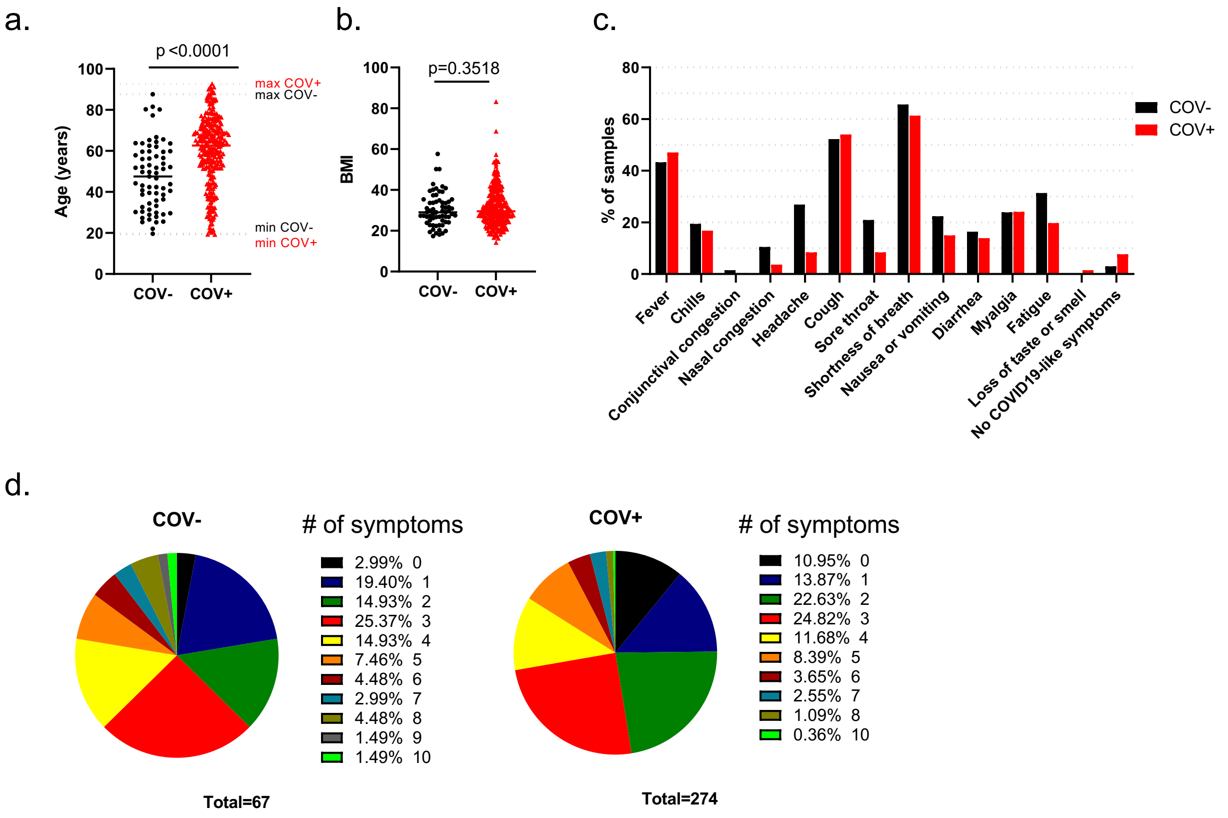

**Figure S1. Age, BMI, and symptom characteristics of the patient cohort.** a) Age distribution of SARS-CoV-2-negative (COV-) and SARS-CoV-2-positive (COV+) patients, including the age span indicated as minimum (min) and maximum (max) age for each group. b) BMI distribution of both COV- and COV+ groups. c) Percentage of COVID-19-related symptoms in both the COV- and COV+ groups. d) Breakdown of the number of COVID-19-related symptoms reported per individual in the COV- and COV+ groups.

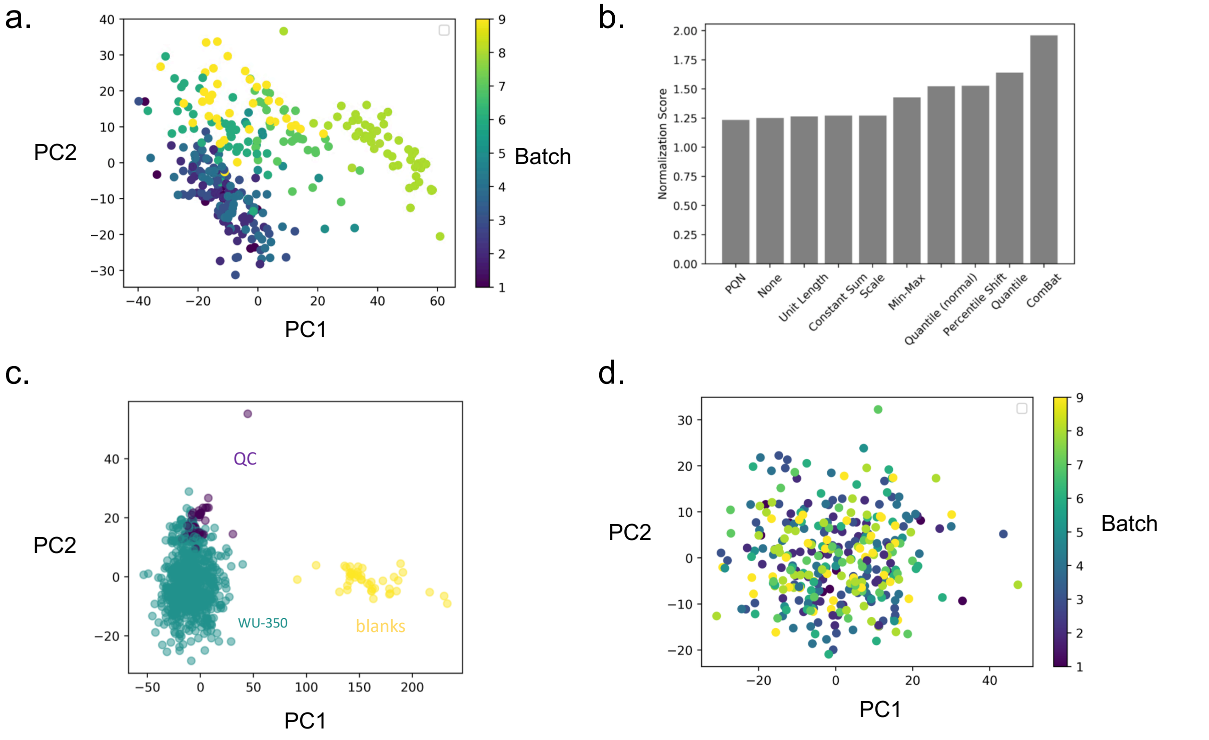

**Figure S2. Batch effects and normalization.** a) Principal component analysis (PCA) plot of metabolic profiles of all research (WU-350), plasma quality control (QC), and blank samples shows show strong batch effects. The color of each dot represents the batch the sample is in. b) Comparison of 10 different batch normalization methods. The y-axis shows the variance remaining in the research samples normalized by the QC variance. c) After ComBat normalization, the PCA plot shows samples cluster by origin (WU-350, QC, blank). d) After ComBat normalization, there is no clustering by batch for research samples.

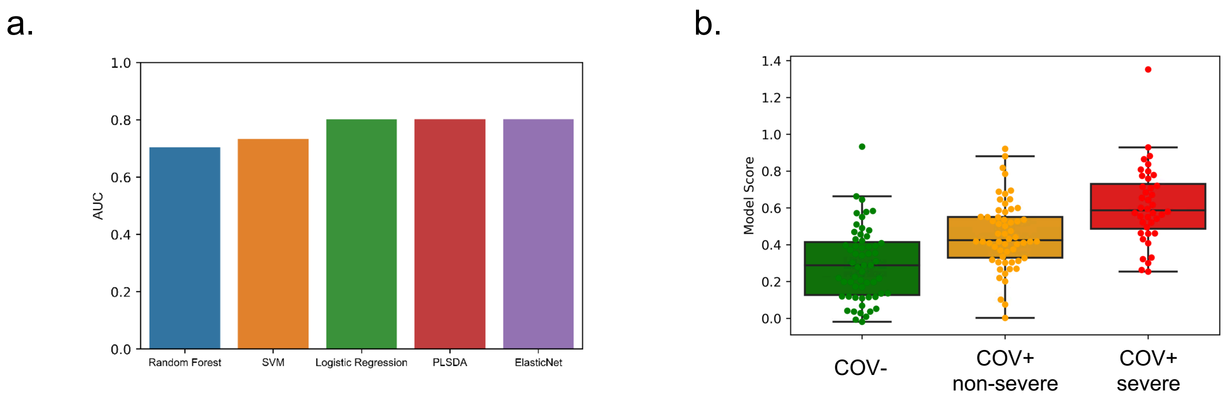

**Figure S3. Model selection and evaluation.** a) 20-fold cross validation was used to select the best modeling framework using the training dataset. The score on the y-axis represents the area under the receiver operating characteristic curve (AUC). An ElasticNet model achieved the highest cross validated performance. b) ElasticNet output when model was trained using all profiled metabolites and the training cohort. The scores for the individuals in the test cohort are broken down by disease status. Box limits represent the quartiles of each sample group. Whiskers are drawn to 1.5x of the inter-quartile range.

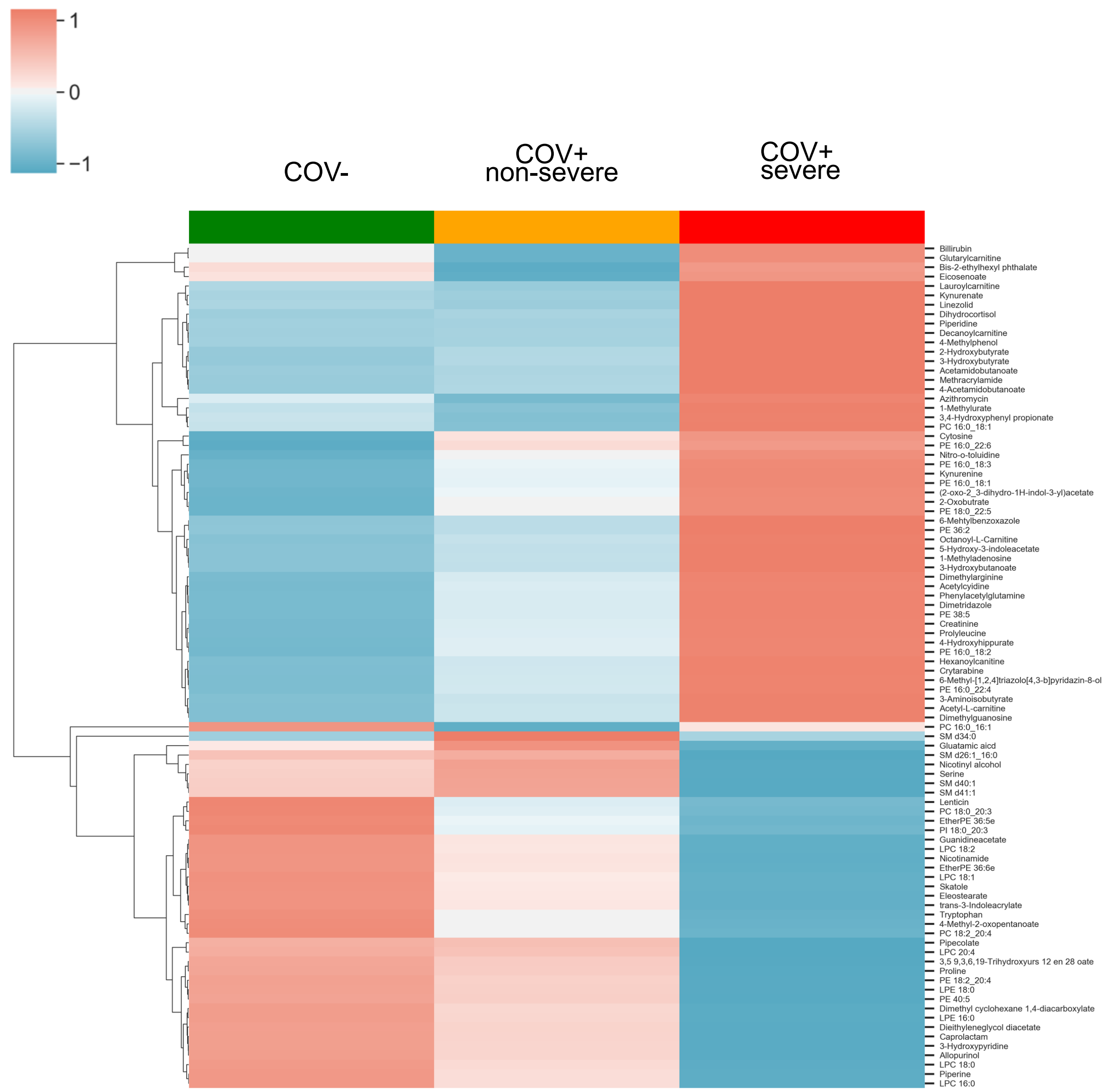

**Figure S4. Clustering of full prediction model metabolites.** Hierarchical clustering analysis of the 93 prediction model metabolites and the mean metabolic profile for COV- (n=67), COV+ non-severe (n=142), and COV+ severe (n=123) patients.

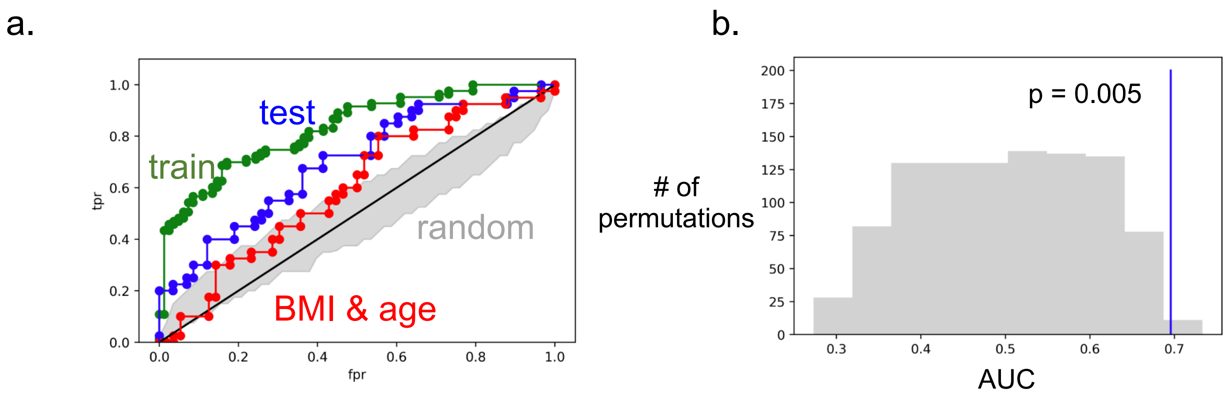

**Figure S5. Model performance using the 25 highly salient metabolites as predictors.** a) Performance on different sets is assessed via ROC curves (x-axis fpr = false positive rate; y-axis tpr = true positive rate) for the training set (green), test set (blue), and baseline performance using only BMI and age as predictors for severe COVID-19 disease (red). Random performance is shown in grey. b) The significance of the model's fit is computed using a permutation test with n=1000 giving an empirical p-value of 0.005.

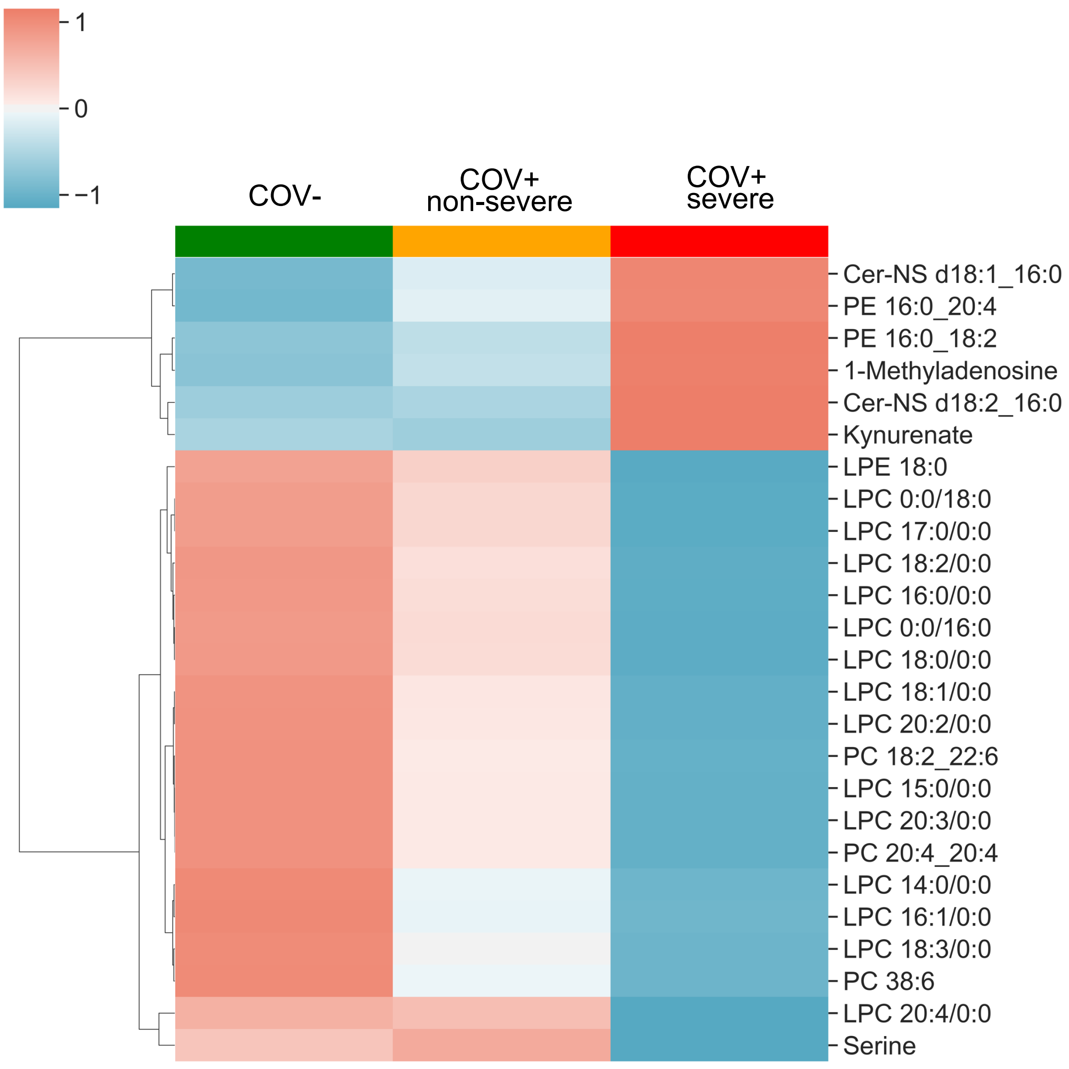

**Figure S6. Clustering of robust predictor metabolites.** Hierarchical clustering analysis of the 25 robust predictor metabolites and the mean metabolic profile for COV- (n=67), COV+ non-severe (n=142), and COV+ severe patients (n=123).

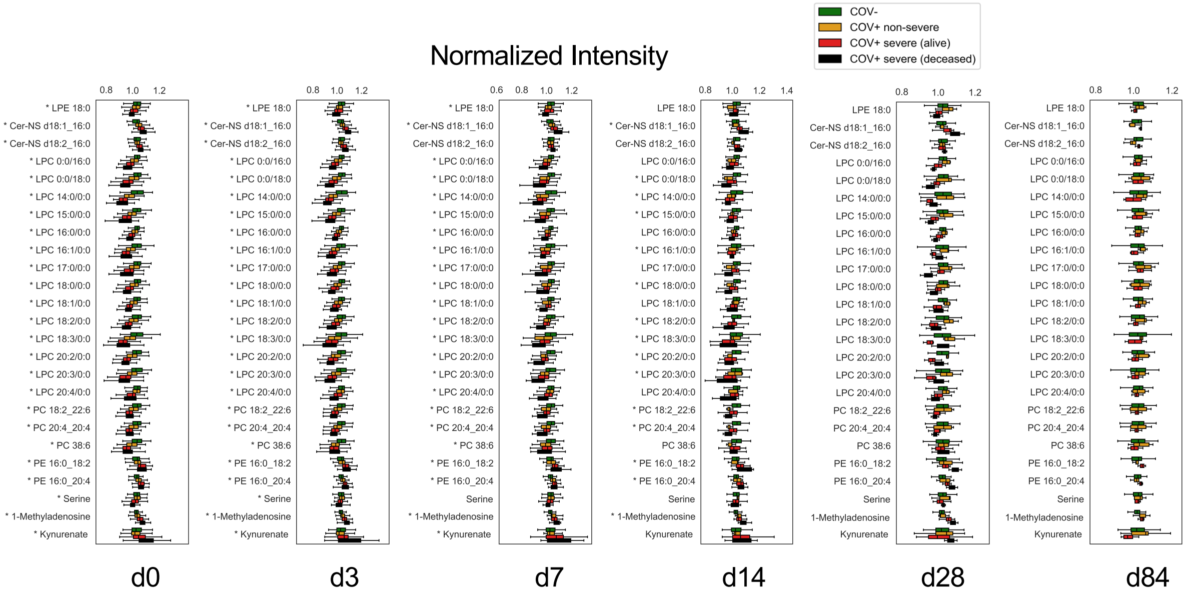

**Figure S7. Comparison of metabolic profiles for deceased and surviving COV+ patients.** Predictor metabolite intensities in COV+ non-severe (orange), surviving COV+ severe patients (red), and deceased COV+ severe patients (black) at each study time point compared to the d0 COV- patients (green). COV+ non-severe patients' metabolite levels recover more quickly than surviving COV+ severe patients. No recovery occurs for deceased COV+ severe patients. * indicates a p-value < 0.05. Statistical significance was assessed using a one-way Welch's ANOVA with Benjamini-Hochberg correction. Box limits represent the quartiles of each sample group. Whiskers are drawn to 1.5x of the inter-quartile range.

**Table S1. Demographics of training cohort.**

| Parameter[1,2] | COV- | COV+ | p-value |
|---|---|---|---|
| N | 38 | 174 | |
| Gender (M/F) | 14/24 | 102/72 | p=0.0145 |
| Age (yr) | 50 ± 18 | 63 ± 16 | p<0.0001 |
| Race (African American/White/other) | 16/22/0 | 132/39/3 | p<0.0001 |
| Body mass index | 30 ± 7 | 30 ± 8 | p = 0.9518 |
| Asymptomatic | 38/0 | 168/6 | p=0.2455 |
| Hospital admission | 14 (36.8%) | 162 (93.1%) | p<0.0001 |
| ICU admission | 6 (15.8%) | 89 (51.1%) | p<0.0001 |
| Intubation and mechanical ventilation | 3 (7.9%) | 42 (24.1%) | p=0.0265 |
| Deceased | 2 (5.2%) | 38 (21.8%) | p=0.0180 |
| Deceased because of COVID-19 | 0 (0%) | 36 (94.7%) | p<0.0001 |

[1] Data are presented as mean ± standard deviation, p-values of numeric parameters calculated using a 2-tailed Student's t-test with unequal variance, p-values of categorical parameters calculated using a chi-square test.

[2] Abbreviations: M – male, F – female, yr – years, B – African American, W – White, O – Other, Y – yes, N – no.

**Table S2. Demographics of test cohort.**

| Parameter[1,2] | COV- | COV+ | p-value |
|---|---|---|---|
| n | 29 | 100 | |
| Gender (M/F) | 14/15 | 56/44 | p=0.5404 |
| Age (yr) | 45 ± 13 | 55 ± 18 | p = 0.0014 |
| Race (African American/White/other) | 16/13/0 | 71/27/2 | p = 0.0785 |
| Body mass index | 31 ± 9 | 34± 11 | p = 0.1784 |
| Symptomatic/Asymptomatic | 27/2 | 85/15 | p=0.2560 |
| Hospital admission | 12 (41.4%) | 91 (90.1%) | p<0.0001 |
| ICU admission | 4 (13.8%) | 40 (40.4%) | p=0.0080 |
| Intubation and mechanical ventilation | 1 (3.4%) | 7 (7.1%) | p=0.4785 |
| Deceased | 2 (6.8%) | 14 (14.1%) | p=0.2995 |
| Deceased because of COVID-19 | 0 (0%) | 12 (85.7%) | p=0.0088 |

[1] Data are presented as mean ± standard deviation, p-values of numeric parameters calculated using a 2-tailed Student's t-test with unequal variance, p-values of categorical parameters calculated using a chi-square test.

[2] Abbreviations: M – male, F – female, yr – years, B – African American, W – White, O – Other, Y – yes, N – no.
